# Mental health services safe staffing: A commissioned rapid scoping review for NHS England

**DOI:** 10.1101/2025.10.23.25338621

**Authors:** Deborah Edwards, Elizabeth Gillen, Nicola Evans, Seren Roberts, Dean Whybrow

## Abstract

This Rapid Scoping Review, commissioned by NHS England, examined recent evidence on safe staffing in mental health services, with a focus on mental health nurses across inpatients and community settings. Drawing on literature published between 2018 and 2024, the review addresses two key questions: the impact of nursing skill mix on patient outcomes and the impact of current deployment models in supporting safe, efficient care in mental health services.

Findings revealed that inadequate staffing and poor skill mix were perceived to compromise quality and safety. Staff shortages were linked to medication errors, incomplete care and increased aggression, while the use of temporary staff and high absence rates were associated with poorer outcomes. Broader literature suggested that increased staffing may reduce suicide-related events, but mental health nurse specific data were limited. Evidence on shift patterns and deployment was also inconclusive.

Overall, the evidence base was fragmented and of low quality, limiting the ability to make definitive policy recommendations. However, the findings may inform future pilot service evaluations and targeted improvements in mental health nurse staffing strategies.

## EXECUTIVE SUMMARY

### What is a Rapid Scoping Review?

This Rapid Scoping Review was completed in four months and aims to explore and summarise available evidence. On the request of the commissioners, quality appraisal was also conducted. It is based on a systematic search of the literature (including grey literature), conducted in January 2024. Priority was given to studies offering robust evidence synthesis, although if these were not identified, primary studies were included. However, due to the volume of evidence retrieved no overarching narrative synthesis was conducted, and the findings should therefore be interpreted with caution.

### Who is this summary for?

NHS England.

### Background / Aim of Rapid Scoping Review

NHS England was commissioned by the National Quality Board to establish a programme to oversee the development of a contemporary suite of improvement resources for safer staffing. The programme aims to provide the NHS in England with a robust, up to date set of resources and guidance which is relevant to current practice and with which NHS boards, NHS managers, staff and patients can be assured and reassured that the decisions they are taking with regards to their workforce continue to be as safe, efficient, effective and sustainable as possible.

The overarching aim of this review was to provide a rapid appraisal of published, international peer-reviewed academic papers and UK policy focused on safe staffing in relation to both inpatient and community mental health services.

The research questions were:

1. What is the current evidence on the impact of mental health nurses’ skill mix across mental health services and patient outcomes?
2. What is the current evidence on the impact of current mental health nurse deployment models to support the provision of safe, efficient patient care across mental health services?

## Results

### Recency of the evidence base

- The previous mental health services evidence review published in 2018 was taken as a starting point.
- This review therefore focussed only on, and included, new published evidence from January 2018 to February 2024 and focused on mental health nurses.

### Extent of the evidence base

- Fifteen relevant primary research studies were found: (observational studies (n=7), modelling studies (n=2), health economics study (n=1), one descriptive study (n=1), qualitative studies (n=2) and mixed methods studies (n=2) from the UK (n=4), Australia (n=3) and one study from each of the following countries the USA, Finland, Japan, Switzerland, Denmark, Greece, Korea, and Italy).
- Four relevant systematic reviews (one low quality [-], two critically low quality [-], one ungraded empty review. The settings where the research was conducted included inpatient (adult n=7; child/ adolescent (n=2), community (n=1), inpatient and community (n=4) and mental health services (n=1)).

## Results

### Question 1: Impact of skill mix models

#### Nursing staff composition

Qualitative evidence revealed that an inadequate skill mix among mental health nurses can negatively impact the safety and quality of mental health care across inpatient and community settings. Review evidence was inconclusive regarding the association between skill mix (the number of registered nurses compared to other groups) and aggression, patient self-harm, the use of restraint and other patient outcomes in inpatient mental health settings, as findings across the studies there were mixed and of low quality. A modelling study found that the presence of a senior nurse leader who provided leadership and support for the whole unit was associated with increased use of seclusion in forensic mental health inpatient units, while having a ward shift co-ordinator who provided leadership and support for each ward was associated with decreased use. However, the ratio of enrolled to registered nurses showed no association with seclusion rates.

#### Staffing levels

Review evidence showed that staff shortages contributed to medication administration errors (critically low-quality evidence). Both qualitative and survey evidence also reveals that understaffing can lead to compromised care including medication errors and certain aspects of nursing care not being completed. Furthermore, qualitative evidence reveals that understaffing can negatively impact mental health care across both inpatient and community settings. In inpatient settings it was felt to lead to increased aggression, compromised patient safety due to poor management of aggression and distress, while adequate staffing was felt to be crucial for ensuring unit safety, quality patient care, and relationship-building.

Review evidence that explored the association between staffing levels in inpatient mental health settings and aggression yielded inconclusive results (low quality evidence). A further review also found mixed findings with both inadequate and higher staffing levels being linked to increased aggression (critically low-quality evidence). Observations of incident reporting data from English inpatient and community mental health settings from 2015 to 2022 showed that there had been a significant rise in incident reporting (although reported incidents of aggression decreased by 7%), and there were no corresponding increases in nurse staffing levels. A cost-effectiveness analysis from the City-128 project favoured scenarios with fewer staff, with higher staffing being consistently correlated with more conflicts. Further quantitative data from child and adolescent psychiatric units reported that predictors of violent incidents included total nursing staff, assistant nurses, patients’ profiles, overall patient count, and the year of the event. Notably, each additional nursing staff member decreased the risk of violence by 60%, while each assistant nurse was associated with a 25% increase in risk.

Review evidence was inconclusive regarding the association between staffing levels in inpatient mental health settings and patient self-harm, the use of seclusion, the use of restraint and a range of other patient outcomes (low-quality evidence). Further quantitative evidence reported no significant associations between the number of permanent staff on a forensic mental health inpatient unit and the use of seclusion, while an increase in registered or enrolled nurses on the same unit was associated with higher seclusion rates. A modelling study found no significant associations between the median number of nurses in hospital settings and relative technical efficiency, which balances resources (e.g., staff) and outcomes (e.g., length of stay or number of patient contacts) compared to similar services. Conversely, in residential non-hospital and outpatient settings, a higher median number of nurses was significantly associated with greater relative technical efficiency.

#### Nurse-bed ratios / Nurse-patient ratios

Review evidence was inconclusive regarding the association between nurse-to-patient ratios in inpatient mental health settings and aggression, patient self-harm, the use of seclusion and the use of restraint, with studies reporting mixed findings (low-quality evidence). In relation to compromised care, lower nurse-patient ratios were correlated with an increased likelihood of medication errors, especially wrong dose administration (critically low-quality evidence).

The findings from quantitative studies showed mixed results regarding nurse-patient ratios and patient outcomes which varied according to the mental health setting. For example, there were no significant associations between the ratio of staff to patients, in a forensic mental health inpatient unit or an adolescent inpatient unit and the use of seclusion. Additionally, there were no significant associations observed for emergency psychiatric treatment involving seclusion and restraint among inpatients. However, a higher number of nurses per 10 beds was associated with an increase likelihood of seclusion and restraint being used. A further observational study reported that higher patient-to-nurse ratios among inpatients were associated with longer hospital stays. However, across all patient categories (inpatients, outpatients, and daycare patients), higher patient-to-nurse ratios were associated with increased hypnotic usage, increased risks of psychiatric readmission within 30 days and a decreased likelihood of a patient receiving emergency psychiatric treatment involving seclusion and restraint.

#### Nurse workforce characteristics

Qualitative evidence emphasises that adequate staffing in mental health settings extends beyond numbers to include staff experience, training, and competence. It was felt that safe staffing is not only about meeting minimum staffing levels but also about ensuring the appropriate distribution of skills and experience with insufficient experience posing risks to care quality across inpatient and community settings. Additionally, a lack of experienced staff or the presence of more junior staff, including new graduates, was perceived as a challenge in managing aggression within inpatient settings.

Review evidence regarding the association between nurses’ gender, years of experience or levels of education and conflict, patient self-harm, use of seclusion, use of restraint and other patient outcomes in inpatient mental health settings was inconclusive as studies showed mixed findings (critically low- and low-quality evidence). Regarding compromised care, further review evidence suggested that junior nurses and newly qualified staff may be more prone to medication errors due to lack of knowledge and increased stress, (critically low-quality evidence).

Other quantitative evidence showed that for each additional male nurse on shift on an inpatient ward, that there was an increased likelihood of mechanical restraint being used. However, in forensic mental health inpatient units, neither the numbers nor the ratio of male to female nursing staff showed significant associations with the use of seclusion. Conversely, within adolescent inpatient units, each additional male nurse on shift was linked to an increased likelihood of seclusion being used, while each additional female nurse was linked to a decreased likelihood. No significant associations were found between nurses’ years of experience or education levels on an inpatient ward and the use of mechanical restraint. Likewise, no significant associations were found between the combined years of mental health experience among staff in an adolescent inpatient unit and the use of seclusion

### Question 2: Deployment models

#### Staff absence

Higher staff absence rates were associated with increased incidents of aggression in inpatient settings (critically low-quality evidence).

#### Use of temporary / agency staff

Qualitative evidence revealed that the presence of agency staff in inpatient settings posed challenges in managing aggression. One review found that high conflict and containment rates were significantly linked to increased levels of unqualified and temporary staff (critically low-quality evidence). Another review found that employing agency staff increased the risk of drug administration errors due to unfamiliarity with processes, medications, and patients (critically low-quality evidence). Other quantitative evidence presented mixed findings regarding agency staff and the use of seclusion. For example, in a forensic mental health unit no significant associations were found between agency staff numbers or permanent-to-casual staff ratios and the use of seclusion. However, in an adolescent inpatient unit the use of seclusion was greater when temporary or agency staff were on shift.

## Discussion

### Staffing levels

Overall, there was a mixed picture about staffing levels related to patient outcomes in both the mental health nursing literature and the findings from the broader literature where mental health nurse data could not be disaggregated from other health personnel. Two broader USA studies reported on suicide related events. Both reported higher rates of suicide related events in areas with staff shortages. One study suggested a reduction in suicide related events where a 1% increase in staffing levels was associated with a 1.6% reduction suicide related events, especially in those areas with the lowest staffing levels. No similar study was identified about mental health nurses or within the UK. Incident data for NHS England reported an increase in incident reporting from 2015-2022, particularly in relation to incidents of self-harm. It was observed that there had been no corresponding growth in nurse staffing levels. In addition, there were survey data that suggested that mental health nurse staff shortages led to more care left undone. Data also showed that more nurses in community settings improved patient outcomes but there was no similar association observed in in-patient settings.

### Skill mix

The broader literature included one study about skill mix in USA veterans’ services that suggested there was no association between patient outcomes and exposure to help from a group of clinicians that included nurses. Within the available mental health nursing qualitative research literature, a balanced distribution of skills and experience was perceived as important. However, systematic review and quantitative findings did not support this view.

### Deployment models

The broader literature consistently reported the negative effects of 12-hour shift patterns compared to 8-hour shifts. While mental health nurses were grouped with other healthcare personnel in this data, no studies included within this review focused on shift patterns for mental health nurses alone.

### Implications for policy

The level of data and consistency of findings is not yet sufficient to make clear policy recommendations about safe staffing of mental health nurses

### Implications for practice

Evidence from the broader literature suggests that increased staffing may lead to a reduction in suicide-related events. While some additional findings related specifically to mental health nursing were identified, the overall picture remains mixed. Current literature may support a pilot service evaluation aimed at increasing staffing in areas with a higher incidence of suicide-related events. Such an evaluation should assess both the potential benefits in those areas and the possible impact on other regions that may experience reduced staffing as a result. This should measure the benefit in those areas but also the costs to areas that may then encounter reduced staffing levels.

### Implications for research

#### Staffing levels

There are significant gaps in knowledge related to safe staffing levels and mental health nursing. Better understanding of decision making about staffing levels is important because staffing may be a deployment as well as a resource issue. USA research demonstrated that increased staffing led to a reduction in suicide related events. Further UK based research is needed to replicate the US study and to disaggregate the finding by professional role, to better inform staffing level decisions.

### Implications for research

#### Skill mix

Further research should explore what nurse staffing works best from the patient perspective, including co-produced recommendations for policy and practice. This should be carried out within the context of different populations and across the life span.

### Implications for research

#### Deployment models

No research explored shift lengths or shift patterns for mental health nurses alone. Further research is indicated that does not interpolate mental health nurses with other heath personnel.

### Implications for research

A fully funded systematic review may offer a more definitive answer to the research questions or broaden the scope. This could explore both data where different professional groups are combined as well as the disaggregated data from available professions, including mental health nursing.

### Conclusions

International evidence regarding hospital nurse staffing in acute care settings suggests that higher levels of registered nurse staffing and a richer skill mix are associated with improved patient outcomes and care quality. In contrast, the evidence base for mental health nursing remains limited and lacks the robustness needed to establish the nature of the relationship between skill mix, nurse staffing levels or ratios, nursing staff composition and key patient outcomes. However, the evidence does suggest a link between quality of care and staffing in mental health settings. Although the review draws on evidence from ten countries, only four studies were conducted in the UK. Given the international variation in nurse education, registration, roles and deployment within mental health services, the applicability of these findings to the UK context should be approached with caution.

## 1. CONTEXT

NHS England was commissioned by the National Quality Board (NQB) to establish a programme to oversee the development of a contemporary suite of improvement resources for safer staffing. The programme aims to provide the NHS in England with a robust, up to date set of resources and guidance which is relevant to current practice and with which NHS boards, NHS managers, staff and patients can be assured and reassured that the decisions they are taking with regards to their workforce continue to be as safe, efficient, effective and sustainable as possible. This has resonance given the extraordinary pressures the NHS workforce endured during the pandemic, and the often significant changes in working practice that this required. The programme will update the existing improvement resources via working groups chaired by strategic influencers and attended by subject matter experts.

A key principle of the NQB Safe and Effective Staffing programme terms of reference is that each setting-specific group (in this case, the Mental Health Services Improvement Resource Professional Reference Group) will use the best available evidence on safe, sustainable staffing models, where it exists, to inform recommendations and the development of their setting-specific improvement.

The overarching aim of this review is to provide a rapid appraisal of published, international peer-reviewed mental health academic papers and UK policy literature that focused on safe staffing in relation to both inpatient and community mental health services. The remit was to build on the previous Mental Health Services evidence review (Lawes et al. 2018). We intended this review to be laser-focused on mental health nurses, the largest professional body within mental health services. This included the skill mix of mental health nurses specifically within nursing teams and across mental health services.

## 2. RESEARCH QUESTION(S)

The commissioning brief set out two areas of interest. The first was to explore the evidence around mental health nursing skill-mix across mental health services focusing on the addition and contribution of other roles and the relationship to patient outcomes. The second area of interest was to investigate to what extent current deployment models support the provision of safe, efficient patient care across mental health services.

Question 1: What is the current evidence on the impact of mental health nurses’ skill mix across mental health services and patient outcomes?

Question 2: What is the current evidence on the impact of current mental health nurse deployment models to support the provision of safe, efficient patient care across mental health services?

## 3. BACKGROUND

### Skill mix models across mental health services

The first question explores the evidence around skill-mix across mental health services focusing on the addition and contribution of other roles and the relationship to patient outcomes. A recent review sought to contextualise skill mix as having three dimensions (Cunningham et al. 2019); specifically 1) mental health nurse role and function, (i.e. skills, abilities, competencies, and knowledge), 2) intra-professional transversality of practice (i.e. grade, ratios of nursing staff, level of qualifications, expertise, experience, education and training), and 3) inter-professional transversality of practice (i.e. ratios of mental health nurses in multi-disciplinary teams). A review conducted by the National Institute for Health and Care Excellence (Rutter et al. 2015) for the Department of Health and NHS England found low quality evidence across 10 studies for the association between inpatient mental health nurse staffing levels and a range of outcomes including conflict and containment rates. The findings of the Mental Health Services evidence review by Lawes et al. (2018) of safe staffing structures agreed with the findings of the Rutter et al. (2015) review, which is that there is limited evidence about optimum staff numbers/ratios and a general lack of research, especially outside of adult mental health inpatient services.

### Deployment models

The second question focuses on investigating to what extent current deployment models support the provision of safe, efficient patient care across mental health services. We operationalised deployment models with the following definition. A deployment model is defined as strategies for deploying mental health nurses within services, for example, covering staff shortfalls by deploying nurses temporarily to unfamiliar wards at short notice (Oliveira et al. 2023). This is important, given that continuity of nursing care with staff that patients are familiar with has been identified as an important characteristic when planning services (NHS Improvement 2018).

## 4. SUMMARY OF THE EVIDENCE BASE

This rapid scoping review was conducted using adapted JBI methodology for scoping reviews (Peters et al. 2020). The protocol is publicly available on Open Science Framework (https://osf.io/9xhrm/). The previous mental health evidence review (Lawes et al. 2018) was taken as a starting point. This new rapid review therefore only included newly published evidence between January 2018 and February 2024. A total of 15 primary research studies met the rapid scoping review inclusion criteria. There were two qualitative studies (Baker et al. 2019; Cranage and Foster 2022) with full details provided in Table 1. There were two mixed methods studies that utilised surveys with both open and closed questions (Delaney et al. 2022; Thompson et al. 2023), and full details are provided in Table 2. There were 11 quantitative studies of which seven were observational retrospective studies that utilised routinely collected data (Fukawsawa et al. 2018; Kodal et al. 2018; Panagiotou et al. 2019; Park et al. 2020; Starace et al. 2018; Woodnutt et al. 2024; Yurtbasi et al. 2021); two were modelling studies (Barr et al. 2022; Diaz-Milanes et al. 2023), one was a descriptive study (Gehri et al. 2023) and one was a health economics study (Kartha and McCrone 2019); full details are provided in Table 3. Additionally, the searches identified four systematic reviews that met the rapid scoping review inclusion criteria (Casey et al. 2023; Ngune et al. 2022; Moyo et al. 2020; Weltens et al. 2021) and full details are provided in Table 4.

**Table 1:**
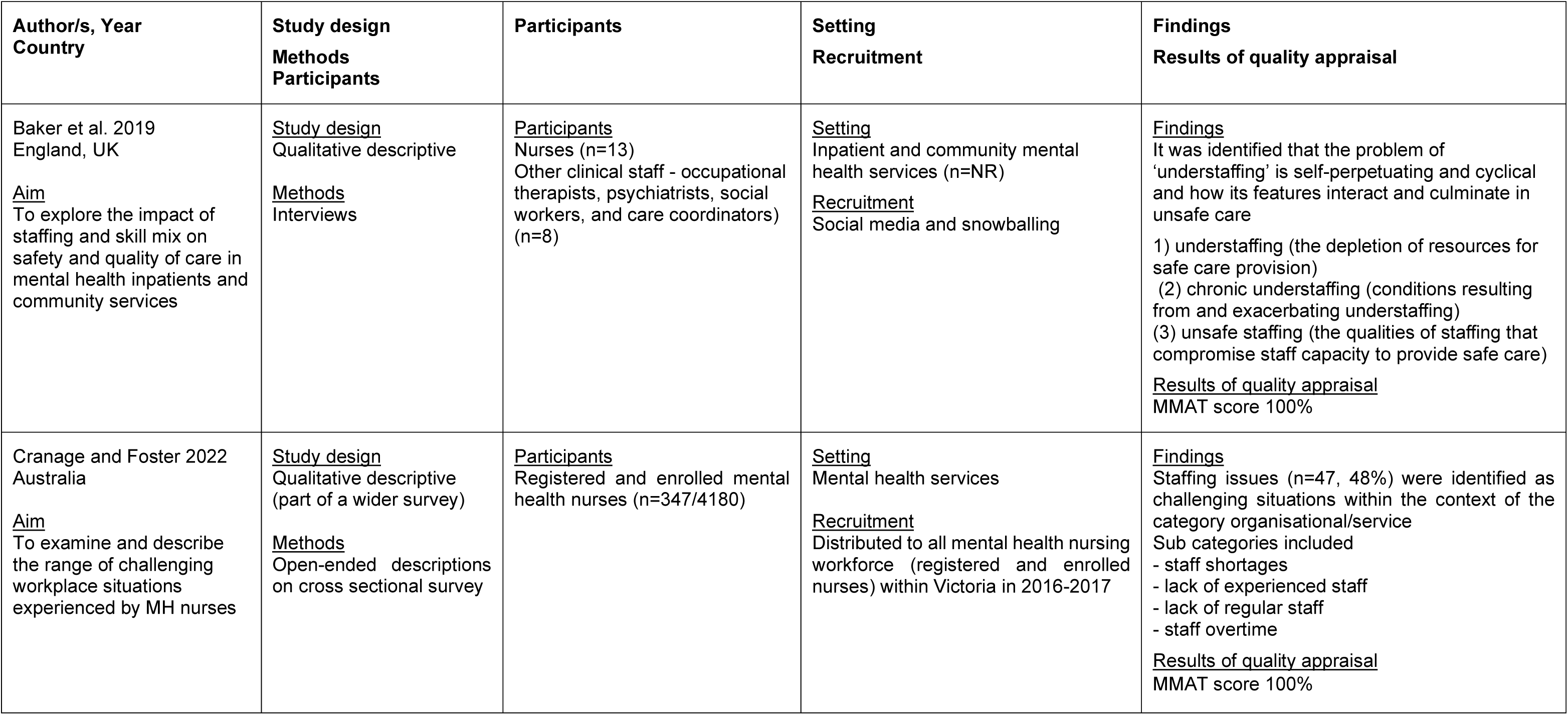
Summary of included primary research evidence from qualitative studies.

**Table 2:** Summary of included primary research evidence from mixed methods studies.

| Author/s, Year<br>Country | Study design<br>Methods<br>Data sources | Participants<br>Setting<br>Recruitment | Outcome/s<br>Outcome measures | Relevant findings<br>Results of quality appraisal |
| --- | --- | --- | --- | --- |
| Delaney et al. 2022<br>USA<br><u>Aim</u><br>To isolate and describe what constitutes quality on inpatient psychiatric units | <u>Study design</u><br>Mixed methods study<br><u>Data sources</u><br>Purposedly developed questionnaire based on 6 dimensions of quality with three open ended responses | <u>Participants</u><br>American Psychiatric Nurse Association members (n=39)<br><u>Setting</u><br>Psychiatric inpatient units<br><u>Recruitment</u><br>Emailed Psychiatric Nurse Association members (n=40) | <u>Outcome/s</u><br>Staff perceptions of quality indicators of inpatient psychiatric care<br><u>Outcome measures</u><br>Survey based on 6 dimensions of quality with open ended responses with a section on adequate staffing levels | <u>Quantitative findings</u><br>Adequate staffing levels - extremely important (82%)<br><u>Qualitative findings</u><br>Participants felt that adequate staffing was more than just numbers, that it should also consider the experience, training and competence level of the staff<br>- that it links to patient safety, staff capacity to spend quality time with patients and to develop needed relationships<br><u>Results of quality appraisal</u><br>MMAT score 60% |
| Thompson et al. 2023<br>UK<br><u>Aim</u><br>To investigate registered mental health nurses' perception of quality of care on their last shift, their self-reported reasons for compromised care and potential impact on patient outcomes | <u>Study design</u><br>Mixed methods study<br><u>Data sources</u><br>2017 Royal College of Nursing employment survey | <u>Participants</u><br>Registered mental health nurses (n=1126)<br><u>Setting</u><br>Mental health inpatient services<br><u>Recruitment</u><br>Secondary analysis of data from the 2017 Royal College of Nursing employment survey<br>No further details provided | <u>Outcome/s</u><br>Nurses' perceived self-reported quality of care delivered, whether they felt it was compromised and the factors that affected quality of care<br><u>Outcome measures</u><br>Likert style questionnaire with open ended responses | <u>Quantitative findings</u><br>34% reported understaffing on their last shift<br>47% reported compromised care on their last shift<br>Top three determinants of compromised care<br>- There were not enough RNs (30.7%)<br>- High patient acuity (29.4%)<br>- Too much time spent on non-nursing duties (27.2%)<br><u>Qualitative findings</u><br>Understaffing is a reason for compromised care and safety for patients and staff<br>Consequences of understaffing<br>- Aggression and injury to self and others<br>-- Minimized or missed care (1:1 time to support and deescalate, omission of escorted leave, opportunities to progress recovery)<br>- Risk of medication errors<br>- Poor management of patient aggression and distress leading to cycles of serious self-harm and incidents<br><u>Results of quality appraisal</u><br>MMAT score 100% |

Key: MH: mental health; MMAT: mixed methods appraisal tool; NR: not reported; RN: registered nurses

**Table 3:** Summary of included primary research evidence from quantitative studies.

| Author/s, Year<br>Country | Study design<br>Methods<br>Data sources | Sample details<br>Setting<br>Recruitment | Outcome/s of interest<br>Outcome measures | Relevant findings |
| --- | --- | --- | --- | --- |
| Barr et al. 2022<br>Australia<br><u>Aim</u><br>To examine whether the use of seclusion in a forensic mental health inpatient setting can be attributed to nursing staff composition and contextual factors | <u>Study design</u><br>Observational<br>Retrospective<br><u>Data sources</u><br>Routinely collected data<br>- Seclusion data<br>- Ward reports<br>- Staff rosters<br>MODELLING STUDY | <u>Sample details</u><br>Seclusion events during 546 shifts<br><u>Setting</u><br>30-bed Forensic mental health inpatient unit (n=1)<br><u>Data collection period</u><br>All shifts over a 6-month period (January to June 2016). | <u>Outcome/s of interest</u><br>Seclusion events<br>Nursing staff composition and contextual factors<br><u>Outcome measures</u><br>Number of seclusion events<br>Number and ratios of:<br>- Males and females<br>- Registrants and enrolled<br>- Temporary and permanent<br>Number of nurses working overtime<br>Staff to patient ratios<br>Roles such as presence of security, lead nurse rostered and permanency of shift coordinator<br>Contextual factors<br>- Day of week, month, shift type<br>- Nursing staff composition<br>- Bed occupancy<br>- Number of admissions<br>- Number of specials | <u>Association between skill-mix and seclusion events</u><br>*Significant Increase in seclusion events*<br>The presence of the hospital senior nurse ($\beta=0.97$; $p=0.18$)<br>*Decrease in seclusion events*<br>The presence of the shift co-ordinator ($\beta -0.84$; $p=0.002$)<br>*No significant differences in seclusion events*<br>Ratio of male:female ($\beta 0.49$; $0.46$)<br>Ratio of perm:casual ($\beta -0.53$ $p=0.64$)<br>Ratio of Enrolled Nurse: Registered Nurse ($\beta -1.18$; $p=0.41$)<br>Number of agency staff ($\beta -0.30$; $p=0.12$)<br>Number of permanent staff ($\beta -0.20$; $p=0.13$)<br><u>Association between nurse-patient ratio and seclusion events</u><br>*No significant differences in seclusion events*<br>Ratio of staff:patient ($\beta -3.09$; $p=0.70$)<br><u>Association between staffing levels and seclusion events</u><br>*Increase in seclusion events*<br>Increased levels of registered nurses ($\beta 0.34$; $p=0.045$)<br>Increased levels of enrolled nurses ($\beta 0.54$; $p=0.014$)<br><u>Association between staffing characteristics and seclusion events</u><br>*No significant differences*<br>Number of male staff ($\beta -0.02$; $p=0.89$)<br>Ratio of male:female ($\beta 0.49$; $p=0.46$)<br><u>Results of quality appraisal</u><br>MMAT score 100% |
| Diaz-Milanes et al. 2023<br>Finland | <u>Study design</u><br>Collective case study | <u>Participants</u> | <u>Outcome/s</u> | <u>Association between staffing levels and relative technical efficacy</u> |
| <u>Aim</u><br>To study the patterns of use and their technical performance (efficiency) of the main types of care of Mental Health services at the Helsinki-Uusimaa region (Finland), and to analyse the potential relationship between technical performance and their corresponding workforce structure | Observational Retrospective<br><u>Data sources</u><br>Routinely collected data<br>REFINEMENT database of Mental Health provisions in Helsinki-Uusimaa<br>MODELLING STUDY | Nurses; Other health personnel<br><u>Setting</u><br>Mental health services (n=229), including inpatient (n=59), residential care (n=92), outpatient care (n=41), and day care (n=37)<br><u>Data collection period</u><br>Secondary data analysis | Relationship between workforce capacity and service technical performance<br><u>Outcome measures</u><br>Relative technical efficiency assessment - the balance between resources (e.g. staff) and outcomes (e.g. length of stay or number of patient contacts) relative to similar service<br>Staff numbers | Hospital acute:<br>Median number of nurses in less (n=12) or more (n=8) efficient services *ns*<br>Residential and non-hospital non-acute settings:<br>Median number of nurses in less (n=0.5) or more (n=4) efficient services (U=9.5; p=0.016)<br>Outpatient and non-acute:<br>Median number of nurses in less (n=3) or more (n=10) efficient services (U=10; p=0.002)<br><u>Results of quality appraisal</u><br>MMAT score 100% |
| Fukawsawa et al. 2018<br>Japan<br><u>Aim</u><br>To clarify the effect of the nurse-to-bed ratio on the use of seclusion and restraint in Japanese general psychiatric wards | <u>Study design</u><br>Observational Retrospective<br><u>Data sources</u><br>Administrative databases<br>Psychiatric Electronic Clinical Observation system | <u>Sample details</u><br>Admissions (n=10,013)<br><u>Setting</u><br>113 general psychiatric wards of 23 institutions<br><u>Data collection period</u><br>All the data for admissions that occurred between April 2015 and March 31, 2017 | <u>Outcome/s of interest</u><br>Associations between nurse-bed ratio and the frequency of use of seclusion and restraint<br><u>Outcome measures</u><br>Number of nurses per 10 beds in each ward<br>Number of admissions exposed to at least one episode of seclusion or mechanical restraint within the first 90 days | <u>Associations between nurse-bed ratio<sup>a</sup> and the frequency of use of seclusion and restraint (mean+SD)</u><br>Admissions without seclusion 4.6±1.2<br>Admissions exposed to seclusion 5.2±1.0<br>p <0.001<br>Admissions without restraint 4.8±1.2<br>Admissions exposed to restraint 5.2±1.0<br>p<0.001<br><u>Associations between nurse-bed ratio and the frequency of use of seclusion and restraint applying multilevel logistic regression analysis</u><br>Seclusion: AOR <sup>a</sup> 2.36 ; 95% CI: 1.55 to 3.60<br>Restraint: AOR 1.74 ; 95% CI: 1.35 to 2.24<br><u>Results of quality appraisal</u><br>MMAT score 100% |
| Gehri et al. 2023<br>Switzerland<br><u>Aim</u><br>To describe the frequency of nursing care left undone in inpatient mental healthcare setting | <u>Study design</u><br>Cross- sectional Prospective (part of a wider study)<br><u>Data sources</u><br>Questionnaires | <u>Participants</u><br>Registered nurses (n=994)<br><u>Setting</u><br>Inpatient units (n=114) across 13 psychiatric hospitals<br><u>Data collection period</u> | <u>Outcomes</u><br>Nursing care left undone<br>Staffing levels<br><u>Outcome Measures</u><br>Nursing care left undone in mental health inpatient setting – 21 item scale; developed for the project<br>Nurse staffing level per shift<br>Staffing and resources adequacy scale | <u>Findings</u><br>The five nursing care activities most often left undone ('often' or 'always')<br>- Evaluating the nursing process (n=272, 30.5%)<br>- Formulating nursing diagnosis (n=225, 27.4%)<br>- Defining care objectives and care plans (n=203, 22.7%)<br>- Reflection of nursing practices on unit (n=204, 21.6%)<br>- Symptom--focused health assessment (n=179, 21.3%) |
| To explore its association with nurse staffing levels |  | Online (n=735) and paper (n=956) surveys distributed by gatekeeper in each of the hospital sites |  | <u>Associations between staffing levels and nursing care left undone</u><br>Shifts with lower-than expected staffing<br>(Estimate: 0.12, 95% CI 0.00 to 0.43)<br><u>Associations between staff reported staffing and resource adequacy and nursing care left undone</u><br>Nurse-reported sufficient staffing and resource adequacy<br>(Estimate: 0.32, 95% CI: -0.40 to -0.24)<br><u>Results of quality appraisal</u><br>MMAT score 100% |
| Kartha and McCrone 2019<br>England, UK<br><u>Aim</u><br>To assess the cost-effectiveness of different staffing levels in relation to conflicts and containments in England, using City-128 study data | <u>Study design</u><br>Health economics study<br>(part of a wider study)<br><u>Data sources</u><br>City-128 data set | <u>Participants</u><br>Day shift nurses (n=29,491)<br>Night shift nurses (n=15,987)<br><u>Setting</u><br>Mental health inpatient care<br>Acute psychiatric wards (n=136) with their patients across 67 hospitals within 26 NHS Trusts<br><u>Data collection period</u><br>Data set from a wider study nor further details reported | <u>Outcome</u><br>Staffing levels<br>Conflict<br>Containment<br>Cost effectiveness<br><u>Outcome measures</u><br>Staff-to-bed ratio<br>- Three scenarios of staffing low, medium and high) bases on shifts in the City-128 dataset<br>Staff numbers<br>Shift type (day-shift / night-shift)<br>Rates of conflict (aggression, rule breaking, alcohol/drug use, absconding, medication refusal and self-harm/suicide)<br>Patient-staff Conflict Checklist<br>Rates of containment (PRN medication, intermittent observation, constant observation, coerced intramuscular medication, show of force, manual restraint, seclusion and time out)<br>The incremental cost and outcomes (total conflict averted and total containment averted) for the day and night groups | <u>Mean costs, total weighted conflicts and containment</u><br>Day shifts (mean cost)<br>£1,102 (low) / £1,360 (medium) / £1,741 (high)<sup>c</sup><br>Mean weighted conflicts<br>14 (low) / 6.55 (medium) / 17.63 (high)<br>Mean weighted containment<br>18.37 (low) / 17.40 (medium) / 20.06 (high)<br>Night shifts (mean cost)<br>£747 (low) / £978 (medium) / £1,381 (high)<br>Mean weighted conflicts<br>11.33 (low) / 12.39 (medium) / 16.23 (high)<br>Mean weighted containment<br>16.25 (low) / 17.25 (medium) / 23.43 (high)<br><u>Cost effectiveness</u><br>High staff group is not cost-effective (because of low effectiveness and high costs) compared to the medium staff group for both total conflicts averted and total containment averted for the day and night groups<br>Medium staff group also is not cost-effective compared to the low staff group<br>Low staff group is the most cost-effective option for the day and night groups<br>The high staff group was also compared with the low staff group and this did not change the cost-effectiveness of the low staff group<br><u>Production function analysis - conflict</u> |
|  |  |  |  | Increased staff numbers were associated with increased numbers of conflicts on shifts, both during the day and night<br>The inclusion of severity measures and the Trust variable did not have any major impact on the coefficients<br><u>Production function analysis – containment</u><br>Increased staff numbers were associated with increased numbers of containment on shifts, both during the day and night<br>The inclusion of severity measures and the Trust variable did not have any major impact on the coefficients<br><u>Results of quality appraisal</u><br>MMAT score 100% |
| Kodal et al. 2018<br>Denmark<br><u>Aim</u><br>To analyse the associations between the incidence of Mechanical Restraint and staff level, staff demographics, patient characteristics and shift of staff | <u>Study design</u><br>Observational<br>Retrospective<br><u>Data sources</u><br>Routinely collected data<br>Administrative databases<br>Care workers from each shift were identified using duty roster | <u>Sample details</u><br>114 cases of mechanical restraint in hospitalised patients with anxiety, depression, bipolar disorders and personality disorders aged 18–67 years<br><u>Setting</u><br>The mood disorder unit, at a psychiatric inpatient department<br><u>Data collection period</u><br>Admission to the psychiatric inpatient department, in the period 1 July 2013 until 30 June 2014 | <u>Outcome/s of interest</u><br>Mechanical restraint<br>Staffing levels<br>Staff characteristics<br>Shifts and change of shifts<br><u>Outcome measures</u><br>Number of patient restrained (enforced fixation to the bed by use of a leather belt around the waist)<br>Number of care workers<br>Age, gender (male/female), education (psychiatric nurse/psychiatric health care assistant/psychiatric nursing aid/unskilled) and experience of each care worker (defined as years of employment at the specific ward) were registered.<br>Psychiatric nursing aids with a 1-year education (that no longer exists) are being replaced by psychiatric health care assistants with a 3-year education<br>Shifts - day shift: 7 am–3 pm evening shift: 3 pm–11 pm, night shift: 11 pm –7 am | <u>Associations between MR and staffing levels</u><br>(univariate analysis)<br>Total number of staff -<br>(OR 1.11; 95% CI 0.95 to 1.30)<br>Number of psychiatric nurses<br>(OR 1.087; 95% CI 0.900 to 1.314)<br>Number of psychiatric health care assistants<br>(OR 1.019; 95% CI 0.832 to 1.249)<br>Number of psychiatric nursing aids<br>(OR 0.951; 95% CI 0.700 to 1.294)<br><u>Significant association between MR and gender</u><br>Males (univariate analysis)<br>(OR 1.64; 95% CI 1.16 to 2.31)<br>Males (multiple regression)<br>(OR 1.44, 95% CI: 1.01, 2.05)<br><u>Association between MR and years of experience</u><br>(univariate analysis)<br>No significant associations – statistics not reported<br><u>Associations between MR and level of education</u><br>(univariate analysis)<br>No significant associations - statistics not reported |

|  |  |  |  | Results of quality appraisal<br>MMAT score 100% |
| --- | --- | --- | --- | --- |
| Panagiotou et al. 2019<br>Greece<br><u>Aim</u><br>To expand on the existing research database by recording, analysing, and explaining the core factors of violent incidents that occurred in the acute psychiatric unit of the Child Psychiatric Hospital of Attica over a 16-year period | <u>Study design</u><br>Observational<br>Retrospective<br><u>Data sources</u><br>Nursing and medical reports and the unit's census | <u>Sample details</u><br>2390 violent incidents across 1600 days (100 days per year over 16 years)<br><u>Setting</u><br>Inpatient<br>Acute Child and Adolescent Psychiatric Unit (Child Psychiatric Hospital of Attica)<br><u>Data collection period</u><br>100 days per year from 1 September to 10 December, of the last 16 years of the unit's operation (1996–2011) | <u>Outcome/s of interest</u><br>Staffing levels<br>Skill Mix<br>Staff characteristics<br>Inpatient violence<br><u>Outcome measures</u><br>Total number of nursing staff in the shift<br>Male to female nurse ratios<br>Registered to assistant nurses ratios<br>Data on diagnostic category of assailants<br>total number of patients; the time of the incident;<br>the type of assault (physical assault towards nursing staff or other patients, towards self, or towards property);<br>The consequences of the incident (i.e., injury, major injury, or no injury)<br>The type of final intervention applied (i.e., verbal de-escalation techniques, PRN medication, or physical restraint) | <u>Association between number of incidents and independent variables (results of multiple regression)</u><br>Total number of patients<br>(IRR 1.0951, 95% CI 1.0667 to 1.1246)<br>Total number of nursing staff<br>(IRR 0.5998, 95% CI 0.5640 to 0.6377)<br>Patients with forensic profile<br>(IRR 1.1406, 95% CI 1.0737 to 1.21116)<br>Number of assistant nurses<br>(IRR 1.2503, 95% CI 1.1812 to 1.3234)<br>Incidents after 2006<br>(IRR 1.6899, 95% CI 1.4688 to 1.9443)<br><u>Results of quality appraisal</u><br>MMAT score 100% |
| Park et al. 2020<br>Korea<br><u>Aim</u><br>To examine the relationships between nurse staffing level and health outcomes of psychiatric inpatients | <u>Study design</u><br>Observational<br>Retrospective<br><u>Data sources</u><br>National Health Insurance Claims Data | <u>Sample details</u><br>70,136 patients aged 19 years who were inpatients in psychiatric wards for at least two days in 2016<br><u>Setting</u><br>Psychiatric wards from tertiary and general hospitals, psychiatric hospitals, and clinics (n=453)<br><u>Data collection period</u><br>All claims data submitted to the Insurance Review and Assessment Service for psychiatric inpatient | <u>Outcome/s of interest</u><br>Nursing staffing levels<br>Health outcomes<br><u>Outcome measures</u><br>Registered nurse-to-inpatient ratio<br>(the average number of inpatients per day divided by the average number of employed psychiatric nurses per day in each hospital)<br>Registered nurse-to-adjusted inpatient ratio - the average number of adjusted inpatients (the sum of the inpatients, outpatients, and daycare patients) per day divided by the average number of employed psychiatric nurses per day in each hospital<br>Nursing staff-to-adjusted inpatient ratio - the average number of adjusted inpatients (the sum of the inpatients, outpatients, and daycare patients) per day divided by the | <u>Association between nurse staffing levels and LOS</u><br>Having more inpatients per RN was significantly associated with longer LOS<br>($\beta=0.02$, SE 0.00, $p < 0.001$)<br>Having more adjusted inpatients per RN was significantly associated with longer LOS<br>($\beta=0.01$, SE 0.00, $p=0.011$)<br>Having more adjusted inpatients per nursing staff was associated with longer LOS<br>($\beta=0.02$, SE 0.00, $p=0.003$)<br><u>Association between nurse staffing levels and readmission within 30 days</u><br>RN to inpatient ratio<br>(AOR<sup>d</sup> 1.01, 95% CI 1.00 to 1.02)<br>Adjusted inpatients per RN<br>(AOR<sup>d</sup> 1.01, 95% CI 1.00 to 1.02)<br>Adjusted inpatients per nursing staff |
|  |  | care in 2016 and combined it with hospital data from the same year (public access data) | average number of employed psychiatric nursing staff (RNs and nurse assistants) per day in each hospital<br>Length of stay<br>Readmission within 30 days<br>Psychiatric emergency treatment (assessing patients' psychiatric symptoms through emergent interviews followed by the administration of seclusion and restraints to prevent harmful behaviours to themselves or others)<br>Use of injected psycholeptics for chemical restraint<br>Hypnotics use | (AOR<sup>d</sup> 1.02, 95% CI 1.00 to 1.04)<br><u>Association between nurse staffing levels and emergency psychiatric treatment</u><br>RN to inpatient ratio (AOR<sup>d</sup> 0.97, 95% CI 0.93 to 1.02)<br>Adjusted inpatients per RN (AOR<sup>d</sup> 0.96, 95% CI 0.92 to 1.00)<br>Adjusted inpatients per nursing staff (AOR<sup>d</sup> 0.92, 95% CI 0.84 to 1.00)<br><u>Association between nurse staffing levels and use of psycholeptics for chemical restraint</u><br>RN to inpatient ratio - no differences ($\beta=0.00$, SE 0.00, $p=0.244$)<br>Adjusted inpatients per RN - no differences ($\beta=0.00$, SE 0.00, $p=0.413$)<br>Adjusted inpatients per nursing staff – no differences ($\beta=0.00$, SE 0.00, $p=0.783$)<br><u>Association between nurse staffing levels risk and use of hypnotics</u><br>RN to inpatient ratio – no differences ($\beta=0.00$, SE 0.00, $p=0.212$)<br>Having more adjusted inpatients per RN was significantly associated with an increased use of hypnotics ($B=0.00$ (rounded off to the second digit after the decimal point, and bigger than 0.00), SE 0.00, $p=0.048$)<br>Having more adjusted inpatients per nursing staff was significantly associated with an increased use of hypnotics ($\beta=0.01$, SE 0.00, $p=0.028$)<br><u>Results of quality appraisal</u><br>MMAT score 100% |
| Starace et al. 2018<br>Italy<br><u>Aim</u><br>Is staffing level is associated with | <u>Study design</u><br>Observational<br>Retrospective<br><u>Data sources</u> | <u>Sample details</u><br>Psychiatric staff<br>Includes mental health nurses<br><u>Setting</u> | <u>Outcome/s of interest</u><br>Staffing levels<br>Antipsychotic prescribing<br><u>Outcome measures</u><br>Total mental health staff (rate per 100,000 inhabitants) | <u>Association between the availability of mental health nurses and the and the number of individuals prescribed antipsychotic drugs</u><br>Univariate analysis: significant inverse association (Kendall's tau -0.485, $p=0.002$) |
| antipsychotic prescribing in community mental healthcare | National mental health information system | Community mental health services<br><u>Data collection</u><br>Based on data collected in 2015 | Mental health nurses (rate per 100,000 inhabitants)<br>Psychiatrists, psychologists, educators & / other staff (rate per 100,000 inhabitants)<br>Antipsychotic prescribing (number of individuals receiving at least one antipsychotic prescription during 2015 per 1,000 inhabitants) | Multivariate analysis:<br>Model 1<sup>e</sup>: ($\beta$ coefficient:-0.792: 95% CI -1.431 to -0.153)<br>Model 2<sup>f</sup>: ($\beta$ coefficient: -0.956: 95% CI -1.770 to -0.142)<br><u>Results of quality appraisal</u><br>MMAT score 100% |
| Woodnutt et al. 2024<br>UK<br><u>Aim</u><br>To compare English national data for incidents and nursing workforce to examine recent trends | <u>Study design</u><br>Observational<br>Retrospective<br><u>Data sources</u><br>Two national datasets of incidents and workforce data for England<br>Incidents – Dataset collated by the National Patient Safety Agency<br>Workforce - National Workforce Dataset | <u>Sample details</u><br>Total reported incidents 51,592 recorded in the first quarter of 2015, and 75,872 reported in the first quarter of 2022.<br><u>Setting</u><br>Inpatient and community mental health settings<br><u>Data collection period</u><br>2015 and 2022 | <u>Outcome/s of interest</u><br>Incident reporting<br>Staffing levels<br><u>Outcome measures</u><br>Self-harm<br><sup>b</sup>Conflict, containment and error<br>Total registered mental health nursing staff<br>Location of staffing<br>- Mental health nurses: registered nurses working within community mental health settings<br>- Registered nurses working in 'other' settings (inpatient or hospital-based mental health settings) | <u>Staffing levels</u><br>6% rise in total registered mental health nursing staff (2015 = 36,543, 2022 = 38,886, difference: 2343)<br><u>Location of services</u><br>Non-community mental health nurses decreased by 12% (2015 = 21,575, 2022 = 19,023)<br>Community mental health nurses increased by 33% (2015 = 14,968, 2022 = 19,863)<br><u>Incident reporting</u><br>46% increase<br>(2015 = 51,592, 2022 = 75,872, difference 24,280)<br><u>Aggression</u><br>7% decrease<br>(2015 = 9085, 2022 = 8449, difference -636)<br><u>Self-harm</u><br>95% increase<br>(2015 = 12,809, 2022 = 25,037, difference 12,228)<br>For every incident of self-harm at the start of the study period, there were 2.85 nurses in employment compared to 1.6 at the end<br><u>Conflict, containment and error<sup>g</sup></u><br>63% increase<br>(2015 = 34,831, 2022 = 56,654)<br>For every incident of conflict, containment or error there were 1.04 nurses in employment at commencement, compared to 0.69<br><u>Results of quality appraisal</u><br>MMAT score 100% |
| Yurtbasi et al. 2021 | <u>Study design</u> | <u>Sample details</u> | <u>Outcome/s</u> | <u>Association between staffing levels and use of seclusion</u> |
| Australia<br><u>Aim</u><br>To examine the relative risk of seclusion occurring in adolescent psychiatric units using both nurse factors and patient factors | Observational Retrospective<br><u>Data sources</u><br>Hospital medical and staff administration records over a 4-year timeframe | 72 afternoon shifts on which seclusion occurred to 216 afternoon shifts<br><u>Setting</u><br>Adolescent inpatient unit<br><u>Data collection period</u><br>Total number of admissions, seclusions, secluded patients, ratio of seclusions to secluded patients, and length of seclusion between 2010 and 2013 | Seclusion<br>Patient factors<br>ICD –10 diagnoses<br>Staff factors<br><u>Outcome measures</u><br>Nurse to patient ratio<br>Total number of nurses<br>Number of male nurses / female nurses<br>Agency/temporary nurses on shift<br>years of mental health experience<br>Total number of inpatients<br>Number of patients with psychotic disorders, depressive disorders, anxiety disorders, other disorders<br>Mean Children's Global Assessment Scale score<br>Total Health of the Nation Outcome Scales<br>Children and Adolescents aggression score<br>Total number of admissions, seclusions , secluded patients<br>Ratio seclusions to patients<br>Average length of seclusion (min)<br>Seclusions (afternoon shift; night shift, (morning shift, crossover) | (univariate analysis)<br>Nurse to patient ratio<br>(OR 6.29, 95% CI 1.27 to 31.15)<br>Total number of nurses<br>(OR 1.24, 95% CI 0.89 to 1.74)<br>Agency / temporary nurses on shift<br>(OR 3.54, 95% CI 1.98 to 6.35)<br><u>Association between staffing characteristics and use of seclusion</u><br>(univariate analysis)<br>Gender (male nurses)<br>(OR 35.68, 95% CI 11.58 to 109.89)<br>Gender (female nurses)<br>(OR 0.27, 95% CI 0.19 to 0.39)<br>Combined years of mental health experience<br>(OR 1.03, 95% CI 1.01 to 1.05)<br>Average years of mental health experience<br>(OR 1.06, 95% CI 0.99 to 1.12)<br><u>Association between patient factors and use of seclusion</u><br>Aggression scores<br>(OR 0.95, 95% CI 0.91 to 0.996)<br>The total number of patients on the unit, diagnosis, and mean Children's Global Assessment Scale score was not associated with risk of seclusion<br><u>Association between staffing characteristics and use of seclusion</u><br>(multivariate analysis)<br>Gender (male nurses)<br>(OR 72.99, 95% CI 13.01 to 409.50)<br>Gender (female nurses)<br>(OR 0.34, 95% CI 0.15 to 0.78)<br><u>Association between staffing levels and use of seclusion</u><br>(multivariate analysis)<br>Nurse to patient ratio<br>(OR 0.90, 95% CI 0.01 to 69.23)<br>Agency / temporary nurses on shift |
|  |  |  |  | (OR 44.37, 95% CI 5.31 to 370.57)<br><u>Association between staffing characteristics and use of seclusion</u><br>(multivariate analysis)<br>Combined years of mental health experience<br>(OR 1.01, 95% CI 0.96 to 1.06)<br><u>Association between patient factors and use of seclusion</u><br>(multivariate analysis)<br>Aggression scores<br>(OR 0.93, 95% CI 0.83 to 1.04)<br><u>Results of quality appraisal</u><br>MMAT score 100% |

Key: AOR: adjusted odds ratio; IRR: incident rate ratio; OIC: Intensive forms of outreach mental health care; MR: mechanical restraint, RN: registered nurse; SD: standard deviation; SE: standard error; VHA: Veterans Health Administration
<sup>a</sup> Adjusted for sex and age and treatment related characteristics included psychiatric diagnosis, form of admission on the first day (voluntary versus involuntary), prescribed dose of antipsychotics at the time of admission (converted into an equivalent dose of chlorpromazine), severity of symptoms, and length of hospital stay.
<sup>b</sup> conflict, containment and error<sup>7</sup> refers to a composite value that includes self-harm, aggression, medication, treatment or procedure, care implementation, documentation, clinical assessment and transfer.
<sup>c</sup> Day Shifts: Low staff scenario has a staff-to-bed ratio of 1:>4; registered staff mean 2.7; non-registered staff mean 1.9; Night shifts: Low staff scenario has a staff-to-bed ratio of 1:>6; registered staff mean 1.5; non-registered staff mean 1.5.
<sup>d</sup> Adjusted for patient characteristics which included age, gender, type of insurance, diagnosis, previous psychiatric hospitalization within the last year, number of psychiatric sub-diagnoses, number of physical sub-diagnoses and Elixhauser Comorbidity Measures score for the last year and system characteristics which included type of hospital, size, ownership, teaching, location, bed operation rate, and RN proportion (the ratio of RNs to total nursing staff).
<sup>e</sup> Model 1 - Adjusted for psychiatric beds (x 100,000 inhabitants); treated prevalence of mental disorders (x 100,000 inhabitants); treated incidence of mental disorders (x 100,000 inhabitants), psychiatric hospital admissions (x 100,000 inhabitants); poverty index; employment rate.
<sup>f</sup> Model 2 - Adjusted for psychiatric beds (x 100,000 inhabitants); treated prevalence of schizophrenia (x 100,000 inhabitants); treated prevalence of bipolar disorder (x 100,000 inhabitants); treated incidence of schizophrenia (x 100,000 inhabitants); treated incidence of bipolar disorder (x 100,000 inhabitants); psychiatric hospital admissions (x 100,000 inhabitants); poverty index; employment rate.
<sup>g</sup> conflict, containment and error<sup>7</sup> refers to a composite value that includes self-harm, aggression, medication, treatment or procedure, care implementation, documentation, clinical assessment and transfer.

**Table 4:** Summary of included review evidence.

| Author/s / Year<br>Aim | Type of review<br>Included studies<br>Dates of search | Setting<br>Focus<br>Outcomes | Findings relevant to the review<br>Results of quality appraisal |
| --- | --- | --- | --- |
| Casey et al. 2023<br>To establish the factors that influence the occurrence of medication administration errors and the reporting of these errors among mental health nurses in mental health hospital settings | <u>Type of review</u><br>Systematic review<br><u>Included studies</u><br>(n=8)<br>-Descriptive including cross-sectional, correlational and longitudinal designs (n=4)<br>-Mixed methods (n=2)<br>-Qualitative (n=2)<br><u>Dates of search</u><br>Not known | <u>Setting</u><br>Inpatient mental health settings<br><u>Focus</u><br>Factors that influence medication administration errors among mental health nurses: Staffing levels; Ward dynamics; Workload<br><u>Outcomes</u><br>Medication administration errors | <u>Findings</u><br>*Work experience or education level*<br>Junior nurses were more prone to medication administration errors (one study)<br>Newly qualified nursing staff described how their lack of knowledge on certain medications and/or patients contributed to errors, which were further compounded by increased feelings of nervousness, stress and pressure to complete tasks (One study).<br>*Staff shortages*<br>Staff shortages contributed to medication administration errors (one study)<br>*Use of agency staff*<br>Use of agency staff led to increased administration error risk due to lack of familiarity with processes, medications, and patients (one study)<br>Inadequate skill mix led to errors and poor decision making when it came to the administration of medicines (one study)<br>*Nurse-patient ratios*<br>Higher patient:nurse ratios were correlated with an increased likelihood of administration errors, especially wrong dose administration (two studies)<br><u>Results of quality appraisal</u><br>Score of 7 out of 11 on the JBI critical appraisal checklist for systematic reviews and research syntheses |
| Moyo et al. 2020<br>To synthesise evidence examining the association | <u>Type of review</u><br>Systematic review<br><u>Included studies</u> | <u>Setting</u><br>Inpatient mental health settings<br><u>Focus</u> | <u>Findings</u><br>*Registered mental health nurse-to-registered nurse ratio* |
| between the mental health-to- registered nurse ratio and patient outcomes (relapse determined by hospital admission) in inpatient mental health settings | Empty review (n=0)<br><u>Dates of search</u><br>Not known | Registered mental health nurse-to-registered nurse ratio<br><u>Outcomes</u><br>Psychiatric readmission (or referral to community crisis services) | No peer-reviewed studies were found that examined the relationship between the ratio of registered mental health staff to registered nurses and psychiatric readmission (or referral to a mental health crisis service) among adult psychiatric inpatients<br><u>Results of quality appraisal</u><br>Score of 6 out of 6 (5 questions not relevant as no included studies found) on the JBI critical appraisal checklist for systematic reviews and research syntheses |
| Ngune et al. 2022<br>To assess the relationship between nursing variables and patient outcomes in acute inpatient mental health settings to determine which outcomes can be used as indicators of the quality of nursing care | <u>Type of review</u><br>Systematic review<br><u>Included studies</u><br>(n=56)<br>- Descriptive including cross-sectional, correlational and longitudinal designs (n=47)<br>- Intervention studies utilising a variety of study designs (n=8)<br>- Economic evaluations (n=1)<br><u>Dates of search</u><br>1995 and 2022 | <u>Setting</u><br>Inpatient mental health settings<br><u>Focus</u><br>Nurse staffing levels<br>Nurse-patient ratios<br>Skill mix (mix of nurse types)<br>Work environment<br>Nurse education<br>Nurse experience<br><u>Outcomes</u><br>Aggression<br>Seclusion<br>Restraint (physical/mechanical or chemical)<br>Absconding<br>PRN medications<br>Special observations<br>Self-harm | <u>Findings</u><br>*Staffing levels, Nurse-patient ratios and skill mix*<br>The results exhibited significant variability across studies, with no consistent pattern emerging in the relationship between staffing levels, nurse-patient ratios, skill mix and patient outcomes (aggression, seclusion, restraint, absconding, PRN medication special observations and self-harm)<br>*Gender*<br>There was inconclusive evidence regarding the impact of nurses' gender and patient outcomes (aggression, seclusion, restraint, absconding, PRN medication special observations and self-harm)<br>*Work experience and education*<br>There was inconclusive evidence regarding the impact of work experience or education on patient outcomes (aggression, seclusion, restraint, absconding, PRN medication special observations and self-harm)<br><u>Results of quality appraisal</u><br>Score of 10 out of 11 on the JBI critical appraisal checklist for systematic reviews and research syntheses |
| Weltens et al. 2021<br>To compile a complete overview of the available knowledge on patient, staff and ward factors that contribute to the | <u>Type of review</u><br>Systematic review<br><u>Included studies</u><br>(n=145)<br>Staff factors (n=55) | <u>Setting</u><br>Inpatient mental health settings<br><u>Focus</u><br>Staff factors: the level of staffing | <u>Findings</u><br>*Staffing levels*<br>Inadequate staffing was associated with increased aggression (three studies)<br>Higher levels of staff was linked to increased aggression (two studies) |
| development of aggression on a general psychiatric admission ward | <u>Dates of search</u><br>January 1999 and December 2019 | <u>Outcomes</u><br>Prevalence of aggressive behaviour | *Use of agency and unqualified and staff*<br>High conflict and containment rates significantly associated with higher levels of unqualified and temporary staff (one study)<br>*Staff absence*<br>Staff being more than average absent from the ward significantly predicted the likelihood of incidents of aggression (one study)<br>*Work experience or education level*<br>No clear effect of work experience or education level on aggression occurrence<br>*Gender*<br>Fourteen studies reported data on the gender of the nurse in relation to aggression<br>No gender differences were found in five studies whereas 9 studies reported that male nurses encountered more aggression<br>*Results of quality appraisal*<br>Score of 9 out of 11 on the JBI critical appraisal checklist for systematic reviews and research syntheses |

The primary research was conducted in UK (n=4), Australia (n=3) and one study from each of the following countries the USA, Finland, Japan, Switzerland, Denmark, Greece, Korea, and Italy.

Nine studies were conducted solely within inpatient settings (seven in adult inpatient settings and two in child and/or adolescent units), one study within community settings and four studies across both inpatient and community settings. A further study described the research as being conducted within mental health services with no further detail provided.

A summary of the primary research evidence is provided below:

- A qualitative study (n=13) that explored the impact of staffing and skill mix on safety and quality of care in mental health inpatients and community services in the UK (Baker et al. 2019).
- An observational retrospective study that used routinely collected data from seclusion events across 546 shifts to model whether the use of seclusion in an Australian forensic mental health inpatient setting can be attributed to nursing staff composition and contextual factors (Barr et al. 2022).
- A qualitative study (as a subset of a larger study) which aimed to examine and describe the range of challenging workplace situations experienced by 347 registered and enrolled mental health nurses in a variety of settings in Australia (Cranage and Foster 2022).
- A mixed methods study that explored the perception of members of the American Psychiatric Nurse Association members (n=39) of quality indicators of psychiatric inpatient care in the USA (Delaney et al. 2022).
- An observational retrospective case study that explored the patterns of use and their technical performance (efficiency) of the main types of care across mental health services (n=229) at the Helsinki-Uusimaa region (Finland); and analysed, through a modelling study, the potential relationship between technical performance and their corresponding workforce structure (Diaz-Milanes et al. 2023).
- An observational retrospective study that sought to clarify the effect of the nurse-to-bed ratio on the use of seclusion and restraint in Japanese general psychiatric wards across 10,013 admissions (Fukawsawa et al. 2018).
- A descriptive cross-sectional study (subset of a larger study with responses from 994 registered nurses) that described the frequency of nursing care left undone in inpatient mental healthcare settings in Switzerland and compared this with nurse staffing levels (Gehri et al. 2023).
- A health economics study (as a subset of a larger study using City-128 study data with data from 29,491 day shift nurses and 15,987 night shift nurses) to assess the cost-effectiveness of different staffing levels in relation to conflicts (aggression, rule breaking, alcohol/drug use, absconding, medication refusal and self-harm/suicide) and containments (PRN medication, intermittent observation, constant observation, coerced intramuscular medication, show of force, manual restraint, seclusion and time out) in England (Kartha and McCrone 2019).
- A retrospective observational study that compared the incidence of mechanical restraint (n=114 cases) with staffing levels, staff demographics, patient characteristics and type of shift (Kodal et al. 2018).
- A retrospective observational study that aimed to record, analyse, and explain the core factors surrounding 2390 violent incidents that occurred across 16 years in an acute psychiatric unit in a hospital in Greece over a 16-year period (Panagiotou et al. 2019).
- A retrospective observational study in inpatient care (70,136 inpatients in psychiatric wards for at least two days in 2016) in Korea that looked at the relationship between nursing ratios to patient outcomes such as length of stay and use of sedation (Park et al. 2020).
- A retrospective observational study looking at the relationship between staffing levels (numbers of staff not reported) and level of antipsychotic prescribing within community mental healthcare in Italy (Starace et al. 2018).
- A mixed methods study of registered mental health nurses’ views (n=1126) on the impact of staffing on quality of care and possible patient outcomes in UK mental health services (Thompson et al. 2023).
- A retrospective observational study that compared English national data for incidents (51,592 recorded in the first quarter of 2015, and 75,872 reported in the first quarter of 2022) defined as patient self-harm, ‘conflict, containment and error with staffing levels across inpatient and community mental health settings (Woodnutt et al. 2024).
- A retrospective observational study in adolescent psychiatric units in Australia looking at the use of seclusion in relation to nurse staffing levels across 72 afternoon shifts (Yurtbasi et al. 2021).

A summary of the review evidence is provided below:

- A systematic review to assess the relationship between nursing variables and patient outcomes in acute inpatient mental health settings to determine which outcomes can be used as indicators of the quality of nursing care. The publications identified in the searches for the review were published between 1995 to 2022 (Ngune et al. 2022 – low quality evidence [-]).
- A systematic review that sought to explore the association between the registered mental health nurse-to-registered general nurse ratio and patient outcomes (relapse determined by hospital admission) in inpatient mental health settings. The date range of the publications identified in the searches for the review was not described (Moyo et al. 2020 – not graded).
- A systematic review to provide an overview of the available knowledge on patient, staff and ward factors that contribute to the development of aggression on a general psychiatric admission ward. The publications identified in the searches for the review were published between 1999 and 2019 (Weltens et al. 2021 – critically low-quality evidence [--]).
- A systematic review to establish the factors that influence the occurrence of medication administration errors and the reporting of these errors among mental health nurses in mental health hospital settings. The date range of the publications identified in the searches for the review was not described (Casey et al. 2023 – critically low-quality evidence [--]).

## 5. KEY FINDINGS

The findings are presented as a series of narrative summaries for each of the two research questions.

### 5.1 Question 1: Impact of skill mix

The evidence regarding the impact of skill mix is presented separately below for nursing staff composition, staffing levels, nurse-bed ratios, nurse-patient ratios, nurse workforce characteristics. The findings are further categorised by the following outcomes: conflict, patient self-harm, use of seclusion, use of restraint, patient safety, compromised care and other patient outcomes alongside the quality score (review evidence) and study design (primary research evidence).

#### 5.1.1 Nursing staff composition

##### Conflict (review evidence)

- The association between skill mix^1^ in inpatient mental health settings and aggression (21 studies) showed considerable variability across all studies included in the review from 1995 to 2022 (Ngune et al. 2022 – low quality evidence [-])

##### Self-harm (review evidence)

- The association between skill mix^2^ in inpatient mental health settings and patient self-harm (six studies) showed considerable variability across all studies included in the review from 1995 to 2022 (Ngune et al. 2022 – low quality evidence [-]).

##### Seclusion (primary research evidence)

- The presence of a senior nurse leader on a forensic mental health inpatient unit was significantly associated with an increase in the use of seclusion (Barr et al 2022 – modelling study).
- The presence of a ward shift co-ordinator^3^ on a forensic mental health inpatient unit was significantly associated with a decrease in the use of seclusion (Barr et al 2022 – modelling study).
- There were no significant associations between the ratio of enrolled to registered nurses on a forensic mental health inpatient unit and the use of seclusion (Barr et al 2022 – modelling study).

##### Patient safety

- Inadequate skill mix can negatively impact the safety and quality of mental health care across inpatient and community settings (Baker et al. 2019 – qualitative evidence).

##### Restraint (review evidence)

- The association between mix of nurse types^4^ in inpatient mental health settings and the use of restraint (17 studies) showed considerable variability across all studies included in the review from 1995 to 2022 (Ngune et al. 2022 – low quality evidence [-]).

##### Other patient outcomes (review evidence)

- The association between mix of nurse types^4^ in inpatient mental health settings and absconding (four studies), PRN medication (two studies), and special observations (three studies) showed considerable variability across all studies included in the review from 1995 to 2022 (Ngune et al. 2022 – low quality evidence [-]).
- The systematic review by Moyo et al. (2020) did not find any studies that investigated the correlation between the ratio of registered mental health nurses to registered general nurses and psychiatric readmission (or referral to a mental health crisis service) among adult psychiatric inpatients.

#### 5.1.2 Staffing levels

##### Compromised care (review evidence)

- A systematic review (dates of search not reported) found one study that reported on staffing levels within inpatient mental health settings and medication errors and found that staff shortages contributed to medication administration errors (Casey et al. 2023 – critically low quality evidence [--]).

##### Compromised care (primary research evidence)

- Understaffing, high patient acuity and spending too much time on non-nursing duties were the three top determinants of compromised care within inpatient settings (Thompson et al. 2023 – survey evidence).
- Lack of time for 1:1 support and de-escalation and the omission of escorted leave were cited as concerning consequences of understaffing within inpatient settings (Thompson et al. 2023 – qualitative evidence).
- When nurses reported that staffing levels were sufficient and that the level of resources were adequate within inpatient settings, then they were significantly more likely to report that less nursing care was left undone (Gehri et al. 2023 – survey evidence).
- There was a significant association between shifts with lower than expected staffing levels within inpatient settings and nursing care left undone (Gehri et al. 2023 – survey evidence).
- Risk of medication errors was cited as a consequence of understaffing within inpatient settings (Thompson et al. 2023 – qualitative evidence).

##### Conflict (review evidence)

- A systematic review that searched for studies from 1999 to 2019 found five studies that investigated the association between staffing levels and aggression. Findings were mixed as both inadequate staffing levels (three studies) and higher staffing levels (two studies) were associated with increased aggression across (Weltens et al. 2021 – critically low quality evidence [--]).
- The association between staffing levels in inpatient mental health settings and aggression showed considerable variability across all studies included in the review from 1995 to 2022 (Ngune et al. 2022 – low quality evidence [-]).

##### Conflict (primary research evidence)

- Observations of incident reporting data in England between 2015 and 2022 within inpatient and community mental health settings found that incident reporting increased significantly, especially with regard to patient self-harm and a composite category of conflict, containment and error^5^ (despite the overall increase in incident reporting, there was a 7% decrease in reported incidents of aggression). The increase in incidents has not been accompanied by a corresponding growth in nurse staffing levels (Woodnutt et al. 2024 - observational study).
- Aggression to self and others (staff and patients) was felt to occur as a result of understaffing within inpatient settings (Thompson et al. 2023 – qualitative evidence).
- Poor management of patient aggression and distress as a result of under staffing were reported to leading to cycles of serious patient self-harm and incidents within inpatient settings (Thompson et al. 2023 – qualitative evidence).
- Cost-effectiveness analysis of the City-128^6^ dataset indicated that in both day and night shifts, a scenario with fewer staff members^7^ proved to be more cost-effective in preventing conflicts even after adjusting for variations inpatient severity (Kartha and McCrone 2019 – health economics study).
- All models generated through a production function analysis consistently demonstrated that higher staffing levels correlated with a rise in conflict occurrences during both daytime and nighttime shift (Kartha and McCrone 2019 – health economics study).
- A multivariate model identified five factors as significant and independent predictors of violent incidents in a child and adolescent psychiatric unit: (i) the total number of nursing staff on duty during the shift; (ii) the number of assistant nurses present during the shift; (iii) the number of patients with social/forensic profiles in the unit; (iv) the overall number of patients in the unit; and (v) the year of the event (post-2006) (Panagiotou et al. 2019 - observational study).
- For each additional member of nursing staff present on a child and adolescent psychiatric unit during a shift there was 60% decrease in the risk of a violent incident. Whereas, for each additional assistant nurse present on a child and adolescent psychiatric unit during a shift there was 25% increase in the risk of a violent incident (Panagiotou et al. 2019 - observational study).

##### Containment (primary research evidence)

- Cost-effectiveness analysis of the City-128 dataset indicated that in both day and night shifts, a scenario with fewer staff members proved to be more cost-effective in containing situations^8^, even after adjusting for variations for acuity of illness (Kartha and McCrone 2019 – health economics study).

##### Patient safety (primary research evidence)

- Adequate and appropriate staffing within inpatient settings was seen as extremely important because it is tied to the safety of the unit, the staff’s ability to dedicate quality time to patients, and the cultivation of relationships (Delaney et al. 2022 – survey and qualitative evidence).
- Poor management of patient aggression and distress due to understaffing was felt to lead to compromised patient safety (Thompson et al. 2023 – qualitative evidence).
- Understaffing can negatively impact the safety and quality of mental health care across inpatient and community settings (Baker et al. 2019 – qualitative evidence).

##### Restraint (review evidence)

- The association between staffing levels in inpatient mental health settings and the use of restraint showed considerable variability across all studies included in the review from in the review from 1995 to 2022 (Ngune et al. 2022 – low quality evidence [-]).

##### Seclusion (review evidence)

- The association between staffing levels in inpatient mental health settings and the use of seclusion showed considerable variability across all studies included in the review from 1995 to 2022 (Ngune et al. 2022 – low quality evidence [-]).

##### Seclusion (primary research evidence)

- There were no significant associations between numbers of permanent staff on a forensic mental health inpatient unit and the use of seclusion (Barr et al 2022 – modelling study).
- Increased numbers of registered or enrolled nurses on a forensic mental health inpatient unit were significantly associated with an increase in the use of seclusion (Barr et al 2022 – modelling study).

##### Self-harm (review evidence)

- The association between staffing levels in inpatient mental health settings and patient self-harm showed considerable variability across all studies included in the review from 1995 to 2022 (Ngune et al. 2022 – low quality evidence [-]).

##### Other patient outcomes (primary research evidence)

- In hospital settings, there were no significant associations between the median number of nurses and relative technical efficiency^9^ (Diaz-Milanes et al. 2023 – modelling study).
- In residential non-hospital and outpatient settings there was a significant association between a higher median number of nurses and greater relative technical efficiency^10^ (Diaz-Milanes et al. 2023 – modelling study).

##### Other patient outcomes (review evidence)

- The association between staffing levels in inpatient mental health settings and absconding, PRN medication, special observations showed considerable variability across all studies included in the review from 1995 to 2022 (Ngune et al. 2022 – low quality evidence [-]).

#### 5.1.3 Nurse-bed ratios

##### Restraint (primary research evidence)

- A higher number of nurses per 10 beds was associated with a 136% increase in the likelihood^11^ of seclusion being used (Fukawsawa et al. 2018 - observational study).

##### Seclusion (primary research evidence)

- A higher number of nurses per 10 beds was associated with a 74% increase in the likelihood^11^ of restraint being used (Fukawsawa et al. 2018 - observational study).

#### 5.1.4 Nurse-patient ratios

##### Conflict (review evidence)

- The association between nurse-to-patient ratios in inpatient mental health settings and aggression showed considerable variability across all studies included in the review from 1995 to 2022 (Ngune et al. 2022 – low quality evidence [-]).
- The association between nurse-to-patient ratios in inpatient mental health settings and patient self-harm showed considerable variability across all studies included in the review from 1995 to 2022 (Ngune et al. 2022 – low quality evidence [-]).

##### Compromised care (review evidence)

- A systematic review (dates of search not reported) found two studies that reported on nurse-to-patient ratios within inpatient mental health settings and medication errors and found higher patient to nurse ratios were correlated with an increased likelihood of administration errors, especially wrong dose administration (Casey et al. 2023 – critically low quality evidence [--]).

##### Seclusion (review evidence)

- The association between nurse-to-patient ratios in inpatient mental health settings and the use of seclusion showed considerable variability across all studies included in the review from 1995 to 2022 (Ngune et al. 2022 – low quality evidence [-]).

##### Seclusion (primary research evidence)

- There were no significant associations between the ratio of staff to patients on a forensic mental health inpatient unit and the use of seclusion (Barr et al 2022 – modelling study).
- There were no significant associations between the numbers of patients (inpatients) per nurse and the risk of psychiatric treatment involving administration of seclusion and restraint (Park et al. 2020 - observational study).
- For each additional patient (inpatients, outpatients, and daycare patients) per nurse there was a 4% decrease in the likelihood of a patient receiving emergency psychiatric treatment involving administration of seclusion and restraint (Park et al. 2020 - observational study).
- For each additional patient (inpatients, outpatients, and daycare patients) per staff (nurses and nursing assistants) there was an 8% decrease in the likelihood of a patient receiving emergency psychiatric treatment involving administration of seclusion and restraint (Park et al. 2020 - observational study).
- There were no significant associations^12^ between the numbers of patients (inpatients only or inpatients, outpatients, and daycare patients) per nurse or staff (nurses and nursing assistants) and the use of injected neuroleptics for chemical restraint (Park et al. 2020 - observational study).
- There were no significant associations between the nurse-to-patient ratio within an adolescent inpatient unit and the use of seclusion (Yurtbasi et al. 2021 - observational study).

##### Restraint (review evidence)

- The association between nurse-to-patient ratios in inpatient mental health settings and the use of restraint showed considerable variability across all studies included in the review from 1995 to 2022 (Ngune et al. 2022 – low quality evidence [-]).

##### Other patient outcomes (primary research evidence)

- Higher numbers of patients (inpatients) per nurse were significantly associated^12^ with longer lengths of psychiatric hospitalisation (Park et al. 2020 - observational study).
- There were no significant associations between the numbers of patients, specifically inpatients per nurse and the use of hypnotics (Park et al. 2020 - observational study).
- Higher numbers across all patient groups (inpatients, outpatients, and daycare patients) per nurse were significantly associated with greater use of hypnotics (Park et al. 2020 - observational study)
- Higher numbers of patients (inpatients, outpatients, and daycare patients) *per staff* (nurses and nursing assistants) were significantly associated^12^ with greater use of hypnotics (Park et al. 2020).
- Higher numbers of patients across all groups (inpatients, outpatients, and daycare patients) per nurse were significantly associated^12^ with longer lengths of psychiatric hospitalisation (Park et al. 2020 - observational study).
- Higher numbers of patients (inpatients, outpatients, and daycare patients) *per staff* (nurses and nursing assistants) were significantly associated^12^ with longer lengths of psychiatric hospitalisation (Park et al. 2020 - observational study).
- For each additional patient (inpatients only) per nurse there was a 1% increased likelihood of psychiatric readmission within 30 days (Park et al. 2020 - observational study).
- For each additional patient across all groups (inpatients, outpatients, and daycare patients) per nurse there was a 1% increased likelihood of psychiatric readmission within 30 days (Park et al. 2020 - observational study).
- For each additional patient across all groups (inpatients, outpatients, and daycare patients) *per staff* (nurses and nursing assistants) there was a 2% increased likelihood of psychiatric readmission within 30 days (Park et al. 2020 - observational study).
- Regions of Italy with higher numbers of mental health nurses have significantly lower rates of individuals prescribed antipsychotic drugs^13^ within community mental health care, even after adjusting for other relevant variables^14^ (Starace et al. 2018-observational study).

#### 5.1.5 Nurse workforce characteristics

##### Patient safety (primary research evidence)

- Adequate staffing within inpatient settings was seen as being more than just numbers in that it should encompass the experience, training, and level of competence of staff (Delaney et al. 2022 - qualitative evidence).
- Safe staffing across community and inpatient services was felt to be not just about having a minimum number of staff it is also about the appropriate distribution of skills and experience (staff time in the role, ward or team and their knowledge about individual patients) (Baker et al. 2019 – qualitative evidence).
- Inadequate experience can negatively impact the safety and quality of mental health care across inpatient and community settings (Baker et al. 2019 – qualitative evidence).

##### Conflict (primary research evidence)

- A lack of experienced staff or the presence of more junior staff, (new graduates) was seen as a challenge when dealing with aggression in inpatient settings (Cranage and Foster 2022 – qualitative evidence).

##### Conflict (review evidence)

- There was inconclusive evidence regarding the association between the gender of nurses working in inpatient mental health settings and patient aggression across all studies examined in the review from 1995 to 2022 (Ngune et al. 2022 – low quality evidence [-]).
- There was inconclusive evidence regarding the association between work experience or levels of education of nurses working in inpatient mental health settings on patient aggression across all studies examined in the review from 1995 to 2022 (Ngune et al. 2022 – low quality evidence [-]).
- A systematic review that searched for studies from 1999 to 2019 reported that there were no clear effects of work experience or education level of nurses working within inpatient mental health settings on patient aggression (Weltens et al. 2021 – critically low-quality evidence [--]).
- A systematic review that searched for studies from 1999 to 2019 found 14 studies that reported on the gender of the nurse working within inpatient mental health settings and patient aggression. There were mixed findings with no gender differences reported across five studies and nine studies reporting that male nurses encountered more aggression (Weltens et al. 2021 – critically low-quality evidence [--]).

##### Self-harm (review evidence)

- There was inconclusive evidence regarding the association between gender of nurses working in inpatient mental health settings and patient self-harm across all studies examined in the review from 1995 to 2022 (Ngune et al. 2022 – low quality evidence [-]).
- There was inconclusive evidence regarding the association between work experience or levels of education of nurses working in inpatient mental health settings and patient self-harm across all studies examined in the review from 1995 to 2022 (Ngune et al. 2022 – low quality evidence [-]).

##### Compromised care (review evidence)

- A systematic review (dates of search not reported) found two studies that explored the experience or level of education of nurses working within inpatient mental health settings. One study found that junior nurses were more prone to medication administration errors and one study found that newly qualified nursing staff described how their lack of knowledge on certain medications and/or patients contributed to errors, which were further compounded by increased feelings of nervousness, stress and pressure to complete tasks (Casey et al. 2023 – critically low quality evidence [--]).

##### Restraint (primary research evidence)

- There were no significant associations between years of experience or levels of education of nurses working on an inpatient ward and the use of mechanical restraint (Kodal et al. 2018 - observational study).
- For each additional male nurse on shift on an inpatient ward, there was a 44% increase in the likelihood of mechanical restraint being used (Kodal et al. 2018 - observational study).

##### Restraint (primary research evidence)

- There was inconclusive evidence regarding the association between the gender of nurses working in inpatient mental health settings and the use of restraint across all studies examined in the review from 1995 to 2022 (Ngune et al. 2022 – low quality evidence [-]).

##### Seclusion (primary research evidence)

- There were no significant associations between the combined years of mental health experience among staff within an adolescent inpatient unit and the use of seclusion (Yurtbasi et al. 2021 - observational study).
- There were no significant associations between the numbers of male nursing staff on a forensic mental health inpatient unit and the use of seclusion (Barr et al 2022 – modelling study).
- There were no significant associations between the ratio of male to female nursing staff on a forensic mental health inpatient unit and the use of seclusion (Barr et al 2022 – modelling study).
- For each additional male nurse on shift within an adolescent inpatient unit, there was a 733% increase in the likelihood of seclusion being used (Yurtbasi et al. 2021 - observational study).
- For each additional female nurse on shift within an adolescent inpatient unit there was a 66% decrease in the likelihood of seclusion being used (Yurtbasi et al. 2021 - observational study).

##### Seclusion (review evidence)

- There was inconclusive evidence regarding the association between the gender of nurses working in inpatient mental health settings and the use of seclusion across all studies examined in the review from 1995 to 2022 (Ngune et al. 2022 – low quality evidence [-]).
- There was inconclusive evidence regarding the association between the work experience or levels of education of nurses working inpatient mental health settings and the use of seclusion across all studies examined in the review from 1995 to 2022 2022 (Ngune et al. 2022 – low quality evidence [-]).

##### Other patient outcomes (review evidence)

- There was inconclusive evidence regarding the association between the gender of nurses working in inpatient mental health settings and patient absconding, PRN medication, special observations across all studies examined in the review from 1995 to 2022 (Ngune et al. 2022 – low quality evidence [-]).
- There was inconclusive evidence regarding the association between the work experience or levels of education of nurses working in inpatient mental health settings and patient absconding, PRN medication, special observations across all studies examined in the review from 1995 to 2022 (Ngune et al. 2022 – low quality evidence [-]).

### 5.2. Question 2: Deployment models

The evidence regarding the impact of optimal deployment models is presented separately below for staff absence and the use of agency/temporary staff. The findings are further categorised by the following outcomes: conflict, seclusion and compromised care alongside the quality score (review evidence) and study design (primary research evidence).

#### 5.2.1 Staff absence

##### Conflict (review evidence)

- A systematic review that searched for studies from 1999 to 2019 found one study that investigated the association between staffing absence in inpatient mental health settings and patient aggression. Staff being absent from the ward more than the average significantly predicted the likelihood of incidents of aggression (Weltens et al. 2021 – critically low-quality evidence [--]).

#### 5.2.2 Use of temporary / agency staff

##### Seclusion (primary research evidence)

- There were no significant associations between numbers of agency staff on a forensic mental health inpatient unit and the use of seclusion (Barr et al 2022 – modelling study).
- For each additional agency or temporary nurse on shift within an adolescent inpatient unit there was a 44% increase in the likelihood of seclusion being used (Yurtbasi et al. 2021 – observational study).
- There were no significant associations between the ratio of permanent to casual staff in a forensic mental health inpatient unit and the use of seclusion (Barr et al 2022 – modelling study).

##### Conflict (review evidence)

- A systematic review that searched for studies from 1999 to 2019 found one study that investigated conflict and containment rates in inpatient mental health settings. High conflict and containment rates were significantly associated with higher levels of unqualified and temporary staff (Weltens et al. 2021–critically low-quality evidence [--]).

##### Conflict (primary research evidence)

- The presence of more agency staff was seen as a challenge when dealing with aggression in inpatient settings (Cranage and Foster 2022 – qualitative evidence).

##### Compromised care (review evidence)

- A systematic review (dates of search not reported) found one study that reported on the use of agency staff and medication errors. The use of agency staff led to increased medication administration error risk due to lack of familiarity with processes, medications, and patients (Casey et al. 2023 – critically low-quality evidence [--]).

## 6. LIMITATIONS OF THE AVAILABLE EVIDENCE

The body of evidence informing this review comes from 10 countries. The countries vary in terms of a) the education and registration of nurses working in mental health care; and, b) the description, definition and allocation of such roles within services. Roles described in services outside the UK may not be directly comparable to those within the UK however there appear to be sufficient commonalities for the evidence to be relevant. Notably, there are differences in the use of restraint: mechanical restraints are not routinely used in the UK, yet their use was reported in several studies from countries such as Japan and Denmark. In this review, the term ‘containment’ was considered to mean seclusion or restraint. We note that self-harm was included in some papers as ‘conflict’ and grouped with externalising aggressive behaviours whereas in the UK, self-harm and suicidal attempts are construed as harm to self, rather than perceived as conflict.

Each individual study was appraised for, quality, with scores reported in Appendix 4. While the primary research studies included high quality non-experimental designs and qualitative descriptive studies, confidence in the findings from the included reviews ranged from low to critically low.

As noted by Lawes et al. (2018), the limited findings across a small number of UK based studies raises concerns about the generalisability of findings, highlighting significant limitations in the available data. A further limitation arises from the types of studies included in this rapid scoping review, specifically, most studies were cross sectional descriptive or qualitative making it difficult to determine causality. However, the included papers offer valuable insights into the impact of skill mix and deployment models on patient outcomes (see section 5).

## 7. DISCUSSION

The overarching aim of this review was to provide a rapid appraisal of published, international peer-reviewed mental health academic papers and UK policy literature that focused on safe staffing in relation to both inpatient and community mental health services. We intended this review to be laser-focused on mental health nurses, the largest professional body within mental health services. This included the skill mix of mental health nurses specifically within nursing teams and across mental health services. The commissioning brief set out two areas of interest. The first was to explore the evidence around mental health nursing skill-mix across mental health services focusing on the addition and contribution of other roles and the relationship to patient outcomes. The second area of interest was to investigate to what extent current deployment models support the provision of safe, efficient patient care across mental health services. To maintain the focus on mental health nursing, we operationalised this into two research questions. Question 1: What is the current evidence on the impact of mental health nurses’ skill mix across mental health services and patient outcomes? Question 2: What is the current evidence on the impact of current mental health nurse deployment models to support the provision of safe, efficient patient care across mental health services?

### 7.1 Summary of the findings: What we found and what we did not

Available evidence on the impact of mental health nurse skill mix focused on nursing staff composition, staffing levels, nurse-bed ratios, nurse-patient ratios, and nurse workforce characteristics. Reported patient outcomes derived from the literature were: conflict, patient self-harm, use of seclusion, use of restraint, patient safety, and compromised care. Inadequate skill mix among mental health nurses negatively impacts safety and quality of care in both inpatient and community settings. However, findings were mixed regarding the association between skill mix and patient outcomes in inpatient mental health settings. Staff shortages were consistently linked to medication administration errors that compromised certain aspects of nursing care. Qualitative evidence highlighted that understaffing contributes to increased aggression and compromised patient safety. Review evidence on the association between staffing levels and aggression in inpatient mental health settings was inconclusive. Adequate staffing extends beyond numbers to include staff experience, training, and competence. The presence of more junior staff, including new graduates, was associated with challenges in managing aggression within inpatient settings. Mixed findings are reported regarding the association between nurses’ gender, years of experience, or education levels and patient outcomes in inpatient mental health settings. No group of studies provided a clear and consistent message about the impact of the mental health nursing team composition on patient outcomes. This may be due to the complexity of healthcare systems, and future research could explore decision making processes around mental health nurse skill mix, which were not well represented in this scoping review.

Evidence on the impact of optimal deployment models focused primarily on staff absence and the use of agency/temporary staff. Associated patient outcomes included conflict, seclusion and compromised care. Higher staff absence rates were associated with increased incidents of aggression in inpatient settings. Mixed findings were observed regarding the association between agency staff and the use of seclusion across different mental health settings. This rapid review did not identify studies examining shift patterns or flexible working arrangements specific to mental health nurses.

### 7.2 Looking beyond mental health nursing

A number of papers were excluded from this review because the findings for mental health nursing could not be disaggregated from other health personnel (see Appendix 3; including reason for exclusion). Those studies represent some of the literature where mental health nurses were interpolated with other groups at either the organisational level (Macro) or interprofessional team level (Meso). To further contextualize this rapid review key findings from relevant excluded studies are presented thematically.

#### 7.2.1 Staffing levels

Cooper et al. (2018) carried out a secondary data analysis in the USA of the association between staffing levels and the adequate provision of either therapy or antidepressants to military veterans with depression. They reported no association between staffing levels and the provision antidepressants or first presentation of depression. There was a small but significant association between staffing and therapy for veterans with recurrent or chronic depression.

McKeown et al. (2019) carried out a qualitative study in the UK about staff and patient perspectives on staffing levels and physical restraint. They reported that forming therapeutic relationships with patients, good communication skills and organisational strategies for reducing the use of restraint was all dependent on sufficient numbers of adequately skilled permanent members of staff. One participant indicated that staff shortages might be due to rostering issues, but this was not reported as a theme.

Ku et al. (2021) carried out a secondary data analysis to test the associations between mental health shortage areas and county-level suicide rates among adults aged 25 and older in the USA from 2010 to 2018. They reported higher suicide rates in areas with greater staff shortages and stated the suicide rates were increasing over time.

Miller et al. (2022) conducted a qualitative study in USA veterans’ services to understand clinicians’ perspectives about the resources necessary to support good functioning mental health treatment teams in the context of low staffing ratios. They reported that combining two smaller teams into one larger team would effectively double up on the number of personnel within the same professional roles, i.e. allow redundancy in professional representation within the team, which meant most disciplines always had somebody available to represent their profession’s perspective in relevant multi-disciplinary meetings.

Feyman et al. (2023) carried out a secondary data analysis in USA veterans’ services to examine the effect of mental health staffing levels on suicide-related events. They reported that a 1% increase in staffing levels was associated with a 1.6% reduction suicide events, especially in those areas with the lowest staffing levels.

#### 7.2.2 Staff skill mix

Boden et al. (2019) carried out a secondary data analysis in USA veterans’ services focused on the association between staffing ratios and treatment access and quality. Total staffing ratios (more clinicians of all types), psychiatrist and therapist staffing ratios were all positively associated with treatment access and quality. While waiting times were important, staffing ratios were more strongly associated with treatment access and quality.

#### 7.2.3 Deployment models

Melathopolous and Cawthorpe (2019) carried out a secondary data analysis of a newly developed Canadian child and adolescent service centralised intake or triage system from 2002 to 2017. They reported an increase in discharge rates, decrease in wait times and length of stay but an increase in staff workload. More specifically an increase in number of tasks and total hours worked but a reduction in time spent per task.

Parker et al. (2023) carried out a qualitative study of peer support and clinical staff in Australia and working in community care units explored perspectives about a new integrated staffing model where peer support workers occupied the majority of roles. Participants reported this model as recovery focused where clinicians provided therapy and support, peer support workers established rapport and applied their lived experience, and residents benefitted from a challenging but enjoyable learning environment.

#### 7.2.4 Shift length

Griffiths et al. (2019) carried out a retrospective longitudinal study in the UK in-patient setting to explore whether 12-hour shifts were associated with a reduction in care hours and staffing costs per patient. The authors reported that when more than 75% of allocated shifts were 12-hour shifts then there was no associated reduction in care hours or costs compared to standard 8-hour shifts. When there was a mixed shift allocation with up to 75% of shifts as 12 hours or longer, this was associated with more care hours per patient per day and increased staffing costs.

Beckman et al. (2022) carried out a retrospective comparative study of USA acute in-patient units (n=32) that used either eight or 12-hour shift patterns. There was a statistically significant difference in favour of 8-hour shift patterns on patient outcomes, measured as challenging behaviours. Most notably, the 12-hour shift group had three times the rate of disruptive events and four times the rate of physical assaults.

(Dall’Ora et al. 2023a) carried out a retrospective longitudinal study using secondary data sources to measure the association between 12-hour shifts and patient incidents in mental health and community hospitals. They reported that 12-hour shifts were associated with increased patient related negative events. More specifically, violence, self-injury and challenging behaviours. There was no association with falls or medication management incidents.

#### 7.2.5 Non-mental health services

When looking beyond mental health services, the international evidence base in relation to general hospital nurse staffing in the acute care setting (Butler et al. 2019; Twigg et al. 2019; Blume et al. 2021; Dall’Ora et al. 2022; Dall’Ora et al. 2023b; Griffiths et al. 2023), there is evidence linking higher registered nurse staffing levels and skill mix to improved patient outcomes and quality. The majority of evidence indicates a reduced risk of death associated with higher nurse staffing levels or skill mix. Additionally, findings indicate reduced complications, such as infections, and shorter lengths of stay, which could significantly contribute to potential cost savings. It is possible that these findings may be transferable to mental health care, but further research is required to establish if this is the case. Equally, a lack of research does not mean that there is necessarily a lack of good practice. Health services may wish to explore exemplars within their organisations in lieu of available research.

#### 7.2.6 Summary of the above findings from section 7.2: Comparing findings from broader studies with those focused on mental health nursing

##### Staffing levels

Overall, there was a mixed picture about staffing levels related to patient outcomes in both the mental health nursing literature and the findings from the broader literature where mental health nurse data could not be disaggregated from other health personnel.

Two broader USA studies reported on suicide related events. Both reported higher rates of suicide related events in areas with staff shortages. One study suggested a reduction in suicide related events where a 1% increase in staffing levels was associated with a 1.6% reduction suicide related events, especially in those areas with the lowest staffing levels. No similar study was identified about mental health nurses or within the UK. However, there was incident data for NHS England that reported an increase in incident reporting from 2015-2022, especially incidents of self-harm. It was observed that there had been no corresponding growth in nurse staffing levels. In addition, there was survey data that suggested that mental health nurse staff shortages led to more care left undone. There was also data that more nurses in community settings improved patient outcomes but no similar association in in-patient settings.

##### Skill mix

The broader literature included one study about skill mix in USA veterans’ services that suggested no association between a group that included nurses and patient outcomes. Within the mental health nursing literature appropriate distribution of skills and experience was perceived as important. However, review and quantitative findings did not support this view.

##### Deployment models

The broader literature consistently reported the negative effects of 12-hour shift patterns compared to 8-hour shifts. While mental health nurses were interpolated within this data, no studies included within this review focused on shift patterns for mental health nurses alone.

### 7.3 Rapid scoping review: Limitations

This rapid scoping review, by its nature included compromises to meet tight timelines and a limited budget. For context, comprehensive systematic reviews require 15-24 months, whereas this rapid scoping review was completed within a three-to-four-month timeframe. While we believe the approach taken helped keep compromises to a minimum, it was still necessary to limit the scope of the review to ensure it remained manageable within the requested timeframe. Consequently, the tender bid and research protocol focused specifically on mental health nurses, excluding studies where mental health nursing data could not be disaggregated from other professional groups. This was approved by the commissioners at the point of application, and the protocol was agreed via correspondence with the stakeholders.

The main methodological compromise involved reducing the use of second reviewer screening from 100% to 10% of potential studies during the study selection process. Despite the rapid timelines, the search terms were well developed, and a wide number of databases were systematically searched. This ensured the review was as comprehensive as possible within the timescale. As is typical with rapid reviews, some relevant studies may have been missed. Despite these limitations, we believe sufficient data was gathered to provide a reasonable overview of current research on mental health nurse skill mix and deployment models, helping to identify potential gaps in knowledge and implications for practice. Please see Section 10 for full details of the methods.

As previously noted, a key limitation of the rapid scoping review was the exclusion of studies where mental health data could not be separated from other professional groups. As a result, while this review adds to knowledge about mental health nursing specifically, it does not include data from studies where mental health nurses was combined with other groups at either the organisational level (Macro) or interprofessional team level (Meso). Given the replicability of this review, further research could be commissioned to explore these aspects by modifying the search times and inclusion and exclusion criteria. Please see section 7.2, where we have contextualised the findings within the wider literature.

### 7.4 Implications for policy

The current level of data and consistency of findings is not yet sufficient to support definitive policy recommendations regarding the safe staffing of mental health nurses. The following implications for practice and research are intended to strengthen the evidence base and guide future policy development.

### 7.5 Implications for practice

There was some evidence in the broader literature suggesting that increased staffing levels may lead to a reduction in suicide related events. While a few additional findings relate specifically to mental health nursing, the picture is more mixed. We consider that further research is needed, particularly studies that account for different professional roles and are focused within the UK context. At a practice level, it may be that the existing literature may be sufficient to support a pilot service evaluation of increasing staffing in areas identified as having higher rates of suicide related incidents. Such an evaluation should assess both the benefits in those areas and the possible costs to other areas that may experience reduced staffing levels as a result.

### 7.6 Implications for research

#### Staffing levels

There are significant gaps in knowledge related to safe staffing levels and mental health nursing. This is partly due to the mixed findings on the relationship between mental health nurse staffing levels and patient outcomes. A deeper understanding of how staffing decisions are made is essential, as staffing may be influenced not only by resource availability but also by deployment strategies. For example, staff may be deployed to manage challenging high-risk situations, yet this may not lead to a reduction in adverse events or better patient outcomes. Such deployment could also impact negatively on personalised care in other areas.

Some evidence from the broader literature suggests that increased staffing may lead to a reduction in suicide related events. While there are relevant findings specific to mental health nursing, the picture remains mixed. Further UK based research is needed, both to replicate existing studies (such as those conducted in the US) and to disaggregate findings by professional role to better inform staffing levels, skill mix and deployment models.

#### Skill mix

The absence of the patient voice in literature concerning staffing levels or the skill mix of mental health nursing staff was notable. Mental health nursing is a unique and privileged role underpinned by developing positive therapeutic relationships with patients. Further research should explore which nurse staffing models are most effective from the patient perspective, ideally through co-producing recommendations for policy and practice. This research should consider different population groups and span different life stages.

#### Deployment models

No research explored shift lengths or shift patterns for mental health nurses alone. However, broader literature indicates that 12-hour shifts in mental health settings may negatively impact patient care. Further research is needed that does not group together mental health nurses with other heath personnel.

Due to the limitations inherent within a rapid review, a fully funded systematic review may offer a more definitive answer to the research questions or broaden the scope. This could explore both data where different professional groups are aggregated as well as disaggregated data from available professions, including mental health nursing.

NHS England has developed research priorities for mental health nursing in the UK (Wadey and Richardson 2024). The above recommendations about safe staffing align to the person-centred practice priorities, specifically 2.2 Policy ambition: preventing suicide and improving support for patients and families, 2.4 Policy ambition: personalised care, and 3.2 Policy ambition: understanding what nurse staffing works best.

## 8. CONCLUSIONS

Overall, the evidence presents a mixed picture on mental health nurses’ skill mix and deployment models in mental health care. Mental health nursing in the UK is relatively unique, as nurses specialise pre-registration. It’s possible that assumptions about the positive impact of mental health nurses are either unsupported by evidence or not directly linked to the unique pre-registration specialism of UK trained nurses. Further UK-based research is needed to explore this issue. We may also need to focus on what different staffing levels enable nurses to do, by measuring or exploring the outcome of specific decision-making interventions or actions carried out by nurses in relation to staffing levels. Finally, a lack of research does not necessarily indicate a lack of good practice. Health services may wish to identify and evaluate exemplars within their organisations in lieu of available robust research and assess their transferability to other settings. However, mental health nursing is an evidence-based profession and healthcare research funders should consider supporting further primary research, as well as systematic or umbrella reviews, related to safe staffing within mental health nursing.

### 8.1. Question 1: Impact of skill mix

- Inadequate skill mix among mental health nurses negatively impacts safety and quality of care in both inpatient and community settings. Mixed findings exist regarding the association between skill mix and patient outcomes in inpatient mental health settings
- Staff shortages contribute to medication administration errors and compromise certain aspects of nursing care.
- Qualitive evidence highlighted that understaffing negatively impacts mental health care, leading to increased aggression and compromised patient safety. However, review evidence regarding the association between staffing levels and aggression in inpatient mental health settings yields inconclusive results.
- Adequate staffing extends beyond numbers to include staff experience, training, and competence.
- The presence of more junior staff, including new graduates, poses challenges in managing aggression within inpatient settings.
- Mixed findings are reported regarding the association between nurses’ gender, years of experience, or education levels and various patient outcomes in inpatient mental health settings.

### 8.2. Question 2: Deployment models

- Higher staff absence rates are associated with increased incidents of aggression in inpatient settings.
- Mixed findings are observed regarding the association between agency staff and the use of seclusion across different mental health settings.

## Data Availability

All data produced in the present study are available upon reasonable request to the authors

## 10. RAPID SCOPING REVIEW METHODS

### Methods

A rapid scoping review was conducted using adapted JBI methodology for scoping reviews (Peters et al. 2020). The protocol is publicly available on Open Science Framework (https://osf.io/9xhrm/). The review is reported according to the Preferred Reporting Items for Systematic Reviews and Meta-analyses extension for scoping reviews (PRISMA ScR) (Tricco et al. 2018).

### Eligibility criteria

The PCC framework was used to inform the eligibility criteria of the initial rapid evidence summary: Population, Concept and Context (Peters et al. 2020).

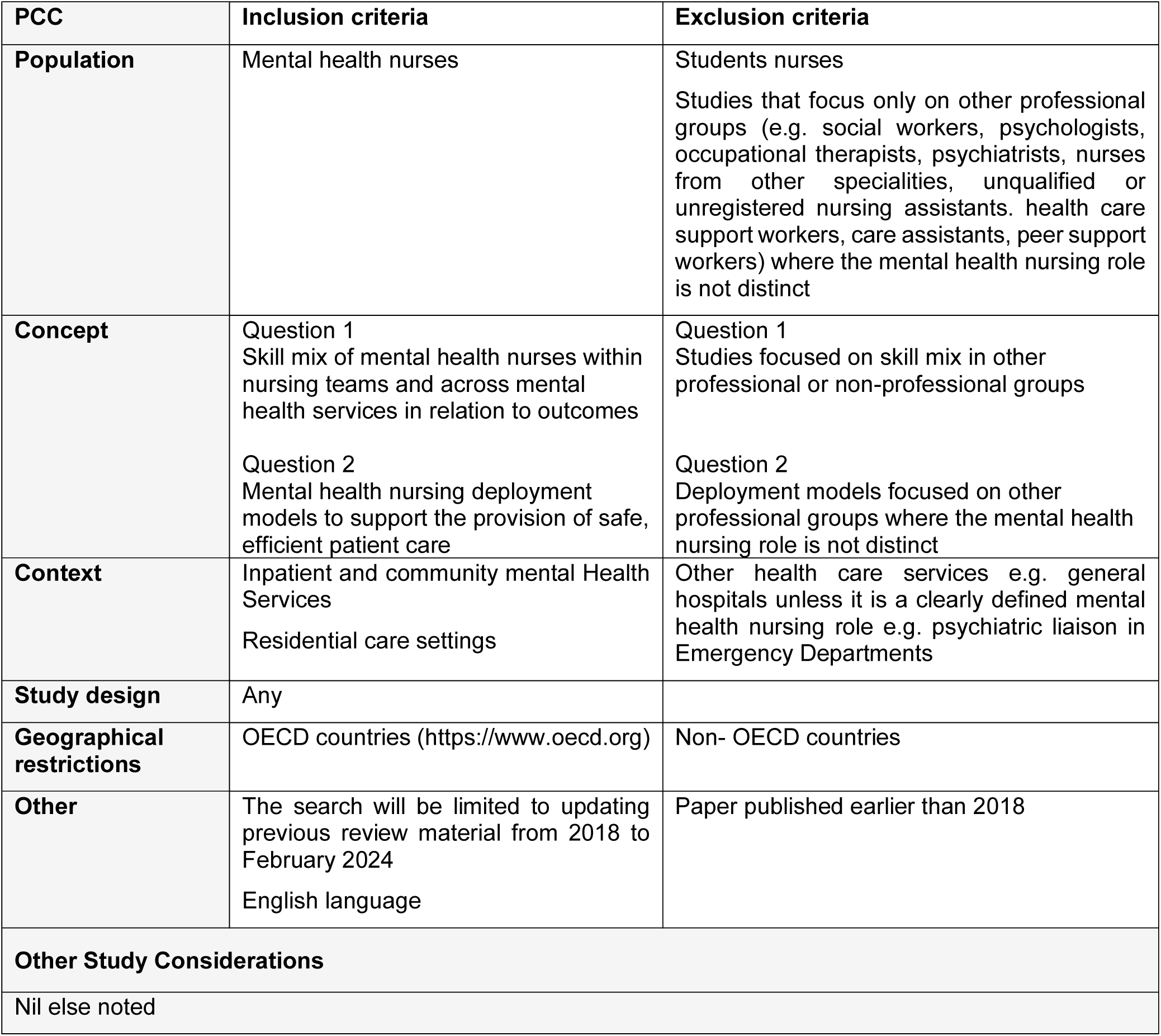

### Literature search

Initial searches of Medline and APA PsycINFO (Ovid platform) were conducted in January 2024 to inform the development of the protocol. The subsequent search results were then used to inform the development of comprehensive search strategies tailored for each information source, for each question.

Comprehensive searches were conducted in February 2024 across seven databases for English language publications from January 2018 to present date:

- On the Ovid Platform: Medline, APA PsycINFO, OVID Emcare, HMIC
- On the EBSCO Platform: CINAHL
- Cochrane (CENTRAL)

The full strategies for each of the databases is presented in Appendix 1.

The websites of key UK third sector and government organisations relevant to the topic area were searched, including: the Royal College of Nursing, Mental Health Nurse Association; Health Education and Improvement Wales (HEIW), NHS England, NHS Wales, NHS Scotland, NHS Northern Ireland, The Health Foundation, National Institute of Health Research (NIHR). No additional research publications were identified.

In a deviation from the protocol we did not conduct forward and backward citation tracking due to time constraints.

### Reference Management

All citations retrieved from the database searches were imported or entered manually into EndNote^TM^ (Thomson Reuters, CA, USA) and duplicates removed. At the end of this process the remaining citations were imported to Rayyan^TM^ and any further duplicates removed.

### Study Selection

All citations were screened by a reviewer from the team, using the information provided in the title and abstract using Rayyan^TM^. A second reviewer from the team screened 10% of these citations with any disagreements resolved through discussion. For citations meeting the inclusion criteria, or in cases in which a definite decision could not be made based on the title and/or abstract alone, the full texts of all citations were retrieved. Each of the full texts were further screened for inclusion by a reviewer from the team, using a purposefully developed screening tool, and all decisions were verified by a second reviewer. Any disagreements were resolved through discussion to reach a consensus. A list of the studies excluded from the review on full text screening can be found in Appendix 2. The flow of citations through each stage of the review process is presented in Appendix 3 in the PRISMA-ScR flow diagram (Tricco et al. 2018).

### Data Extraction

All demographic and outcome data was extracted directly into tables by one reviewer and checked by another. This process was piloted on eight studies. The data extracted includes specific details about the populations, study methods and outcomes of significance to the review questions.

### Assessment of Methodological Quality

Methodological quality was assessed by one reviewer (and judgements verified by a second reviewer). Overall critical appraisal scores are presented in Appendix 4. Systematic reviews were appraising using the JBI critical appraisal checklist for systematic reviews and research syntheses (Aromataris et al. 2015). Where a particular point for inclusion was regarded as “unclear” it was given a score of zero. Where a particular point for inclusion was regarded as “not applicable” this point was taken off the total score. Primary research studies were appraised using the Mixed Methods Appraisal Tool (MMAT-Version 2018) (Hong et al. 2018).

### Overall confidence in the results of reviews

Alternative appraisal tools that can be used for assessing the quality of SRs, evidence maps and overviews of reviews include the AMSTAR-2 (Shea et al. 2017). While in this rapid review, the JBI critical appraisal checklist for systematic reviews and research syntheses (Aromataris et al. 2015) was selected due to its ability to be completed more swiftly than AMSTAR-2, five of the JBI quality checklist questions could be matched to the domains deemed critical in the AMSTAR-2 which were considered relevant to this review.

As a result, the JBI domains considered critical after the mapping include the following:

Q3: Was the search strategy appropriate?

Q4: Were the sources and resources used to search for studies adequate? Q5: Were the criteria for appraising studies appropriate?

Q8: Were the methods used to combine studies appropriate? Q9: Was the likelihood of publication bias assessed?

Each review was then assessed based on the answers provided to the four critical domains as well as the remaining, non-critical, domains, and an overall rating of quality for each review was generated as detailed below.

- High quality [++]: No or one non-critical weakness. The systematic review provides an accurate and comprehensive summary of the results of the available studies that address the question of interest.
- Moderate quality [+]: More than one non-critical weakness^15^ the systematic review has more than one weakness but no critical flaws. It may provide an accurate summary of the results of the available studies that were included in the review.
- Low quality [-]: One critical flaw with or without non-critical weaknesses. The review has a critical flaw and may not provide an accurate and comprehensive summary of the available studies that address the question of interest.
- Critically low [- -]: More than one critical flaw with or without non-critical weaknesses. The review has more than one critical flaw and should not be relied on to provide an accurate and comprehensive summary of the available studies.

### Synthesis

The data has been reported narratively as a series of thematic summaries across each research question (Thomas et al. 2017).

## 11. APPENDICES

**Appendix 1:**
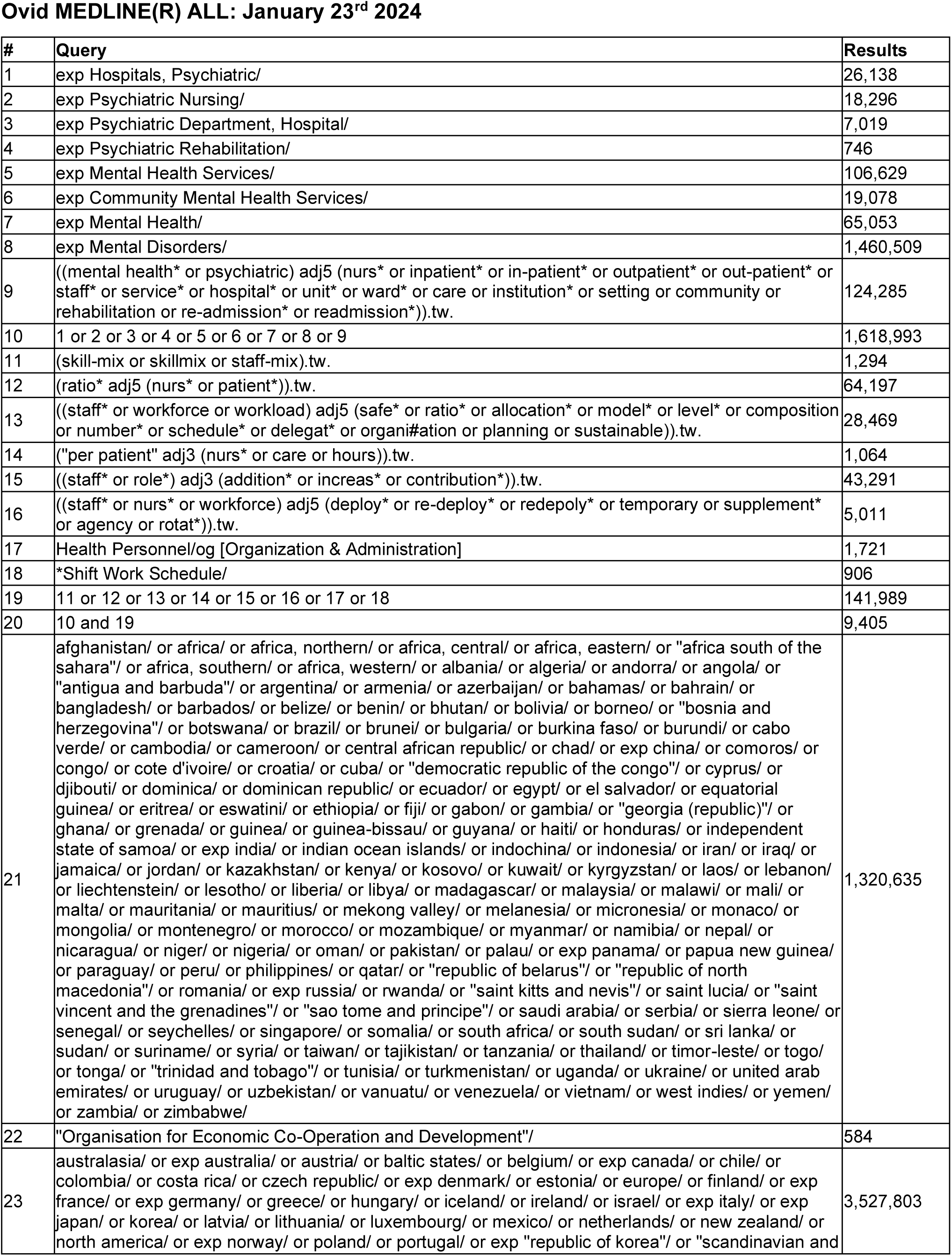
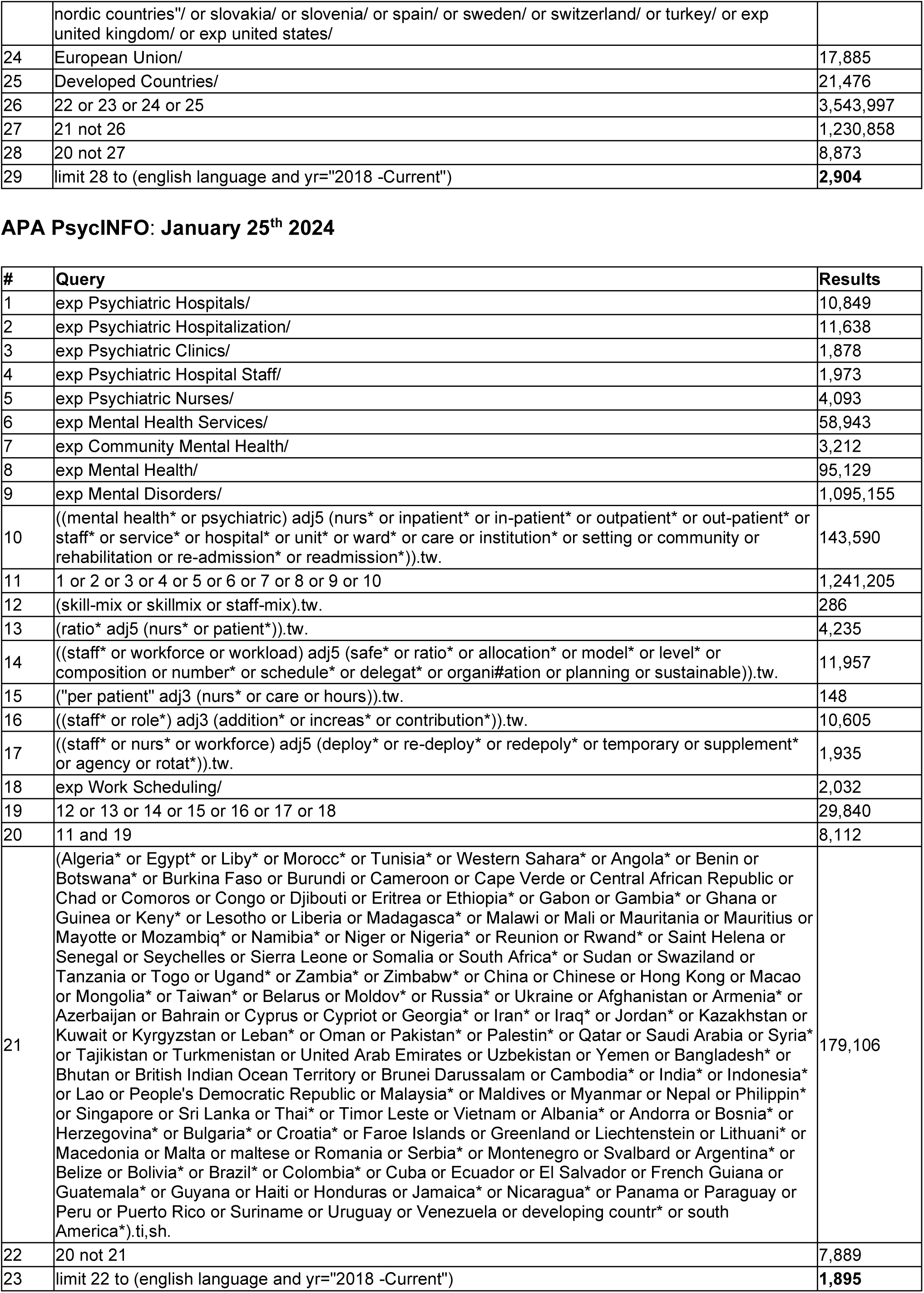
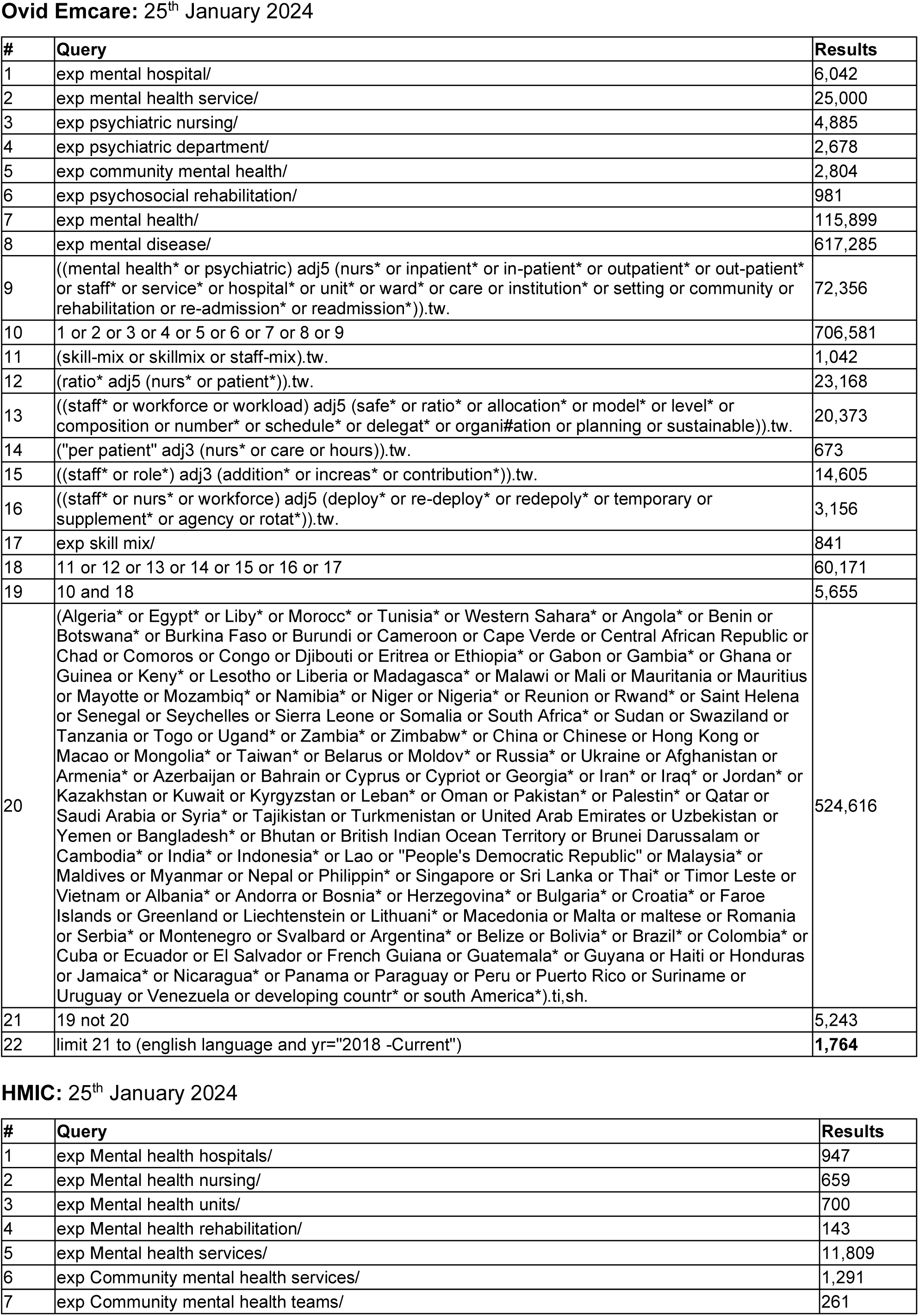
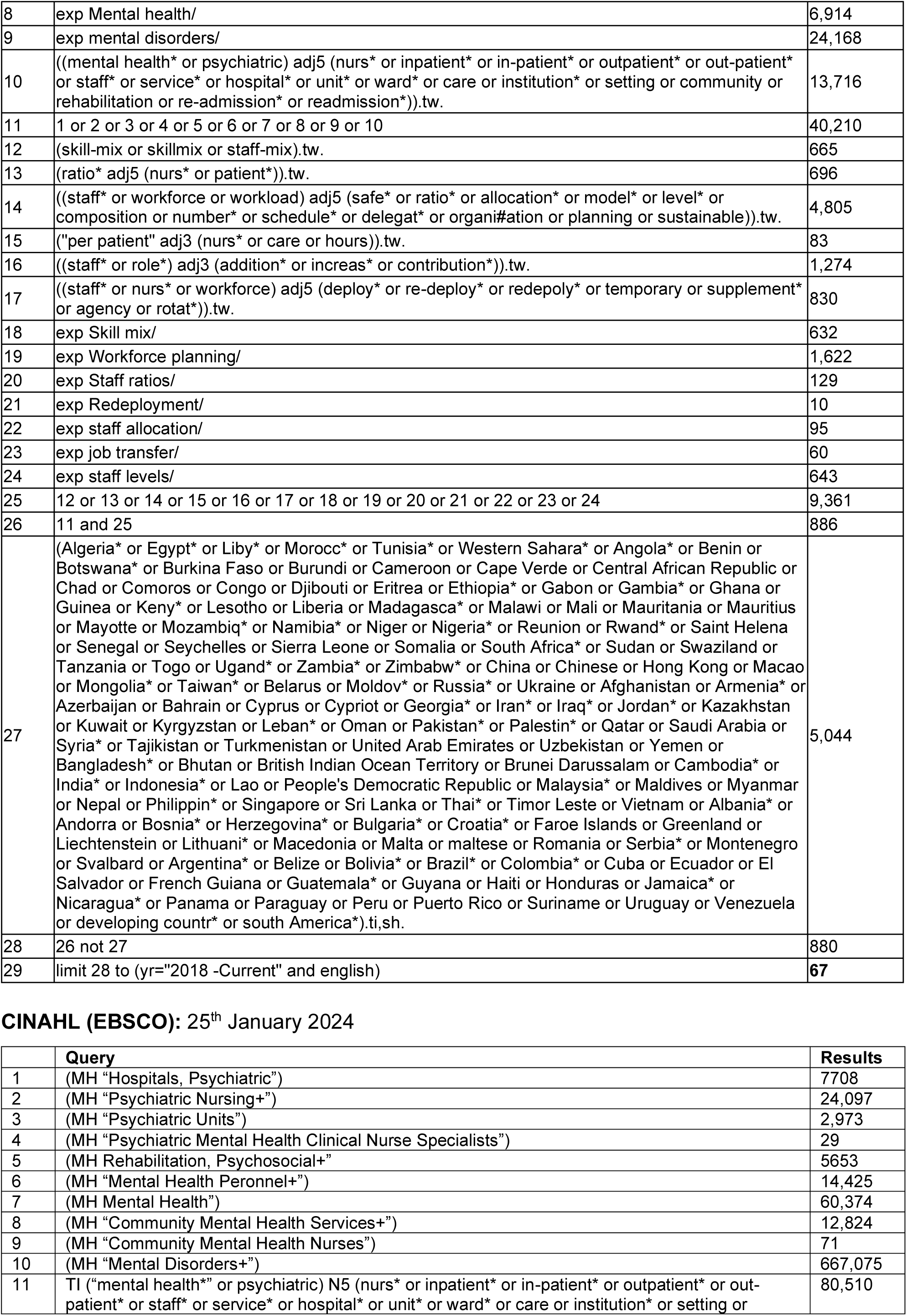
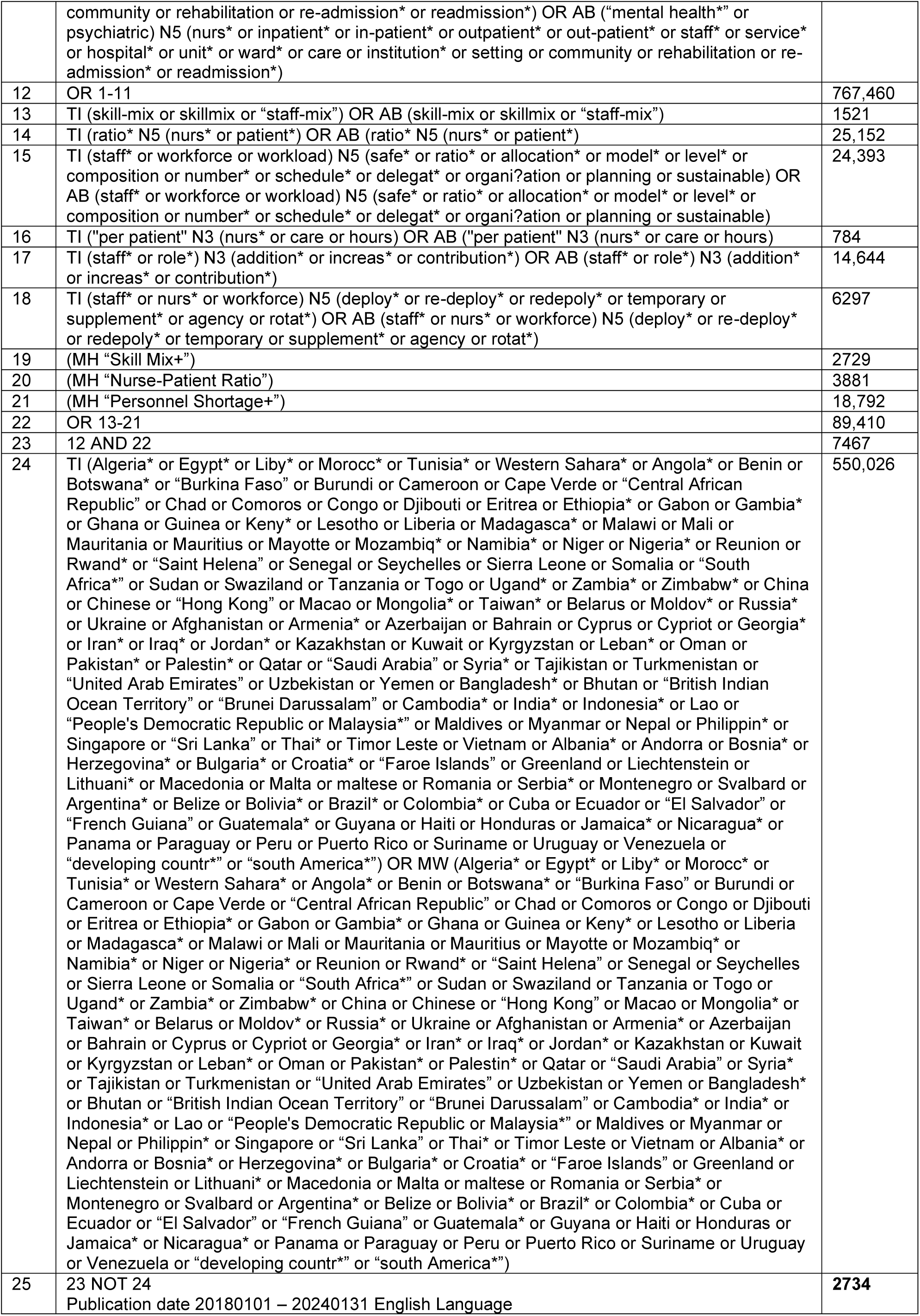
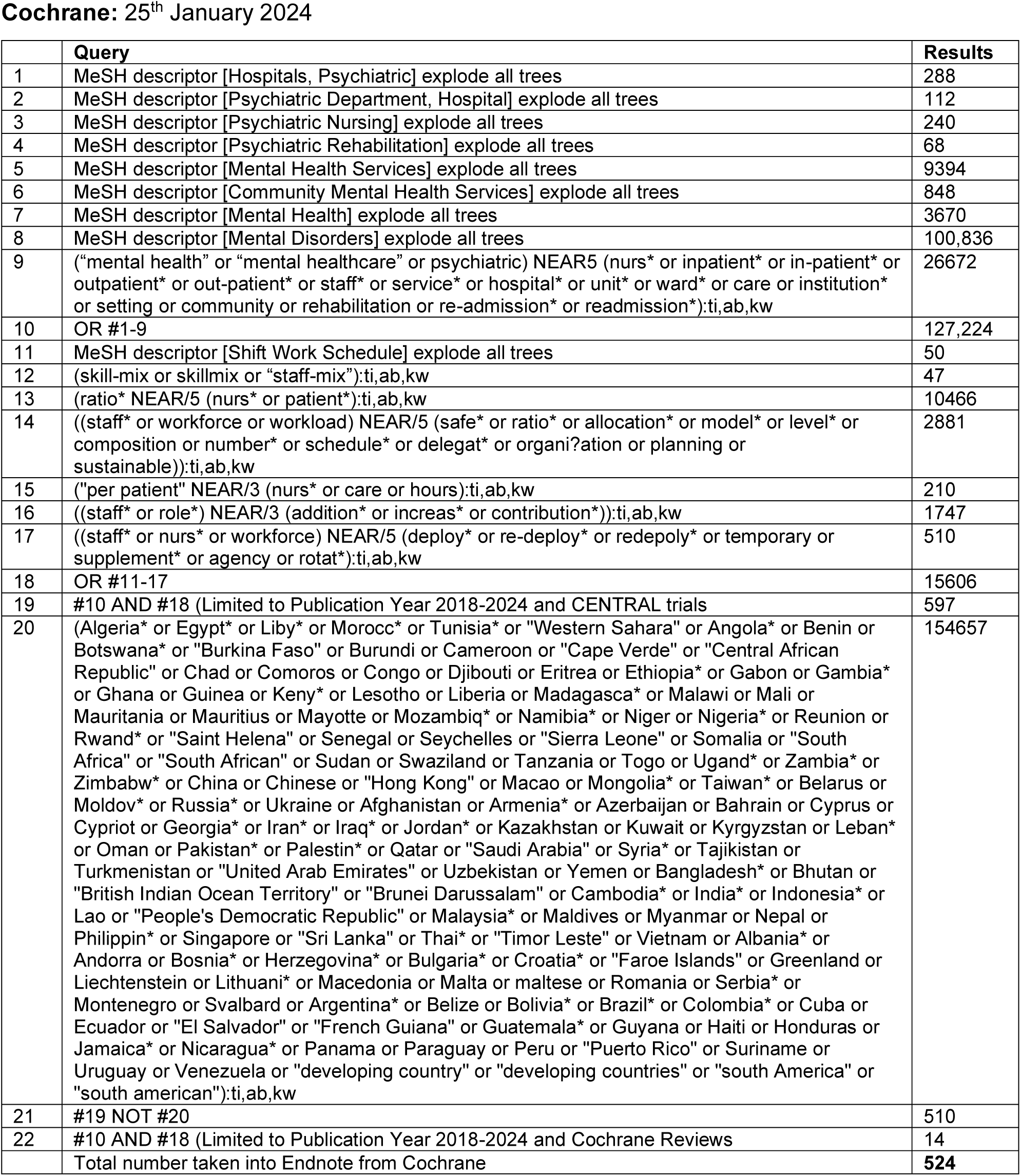
Search strategies.

**Appendix 2:** Final search numbers.

| Database | Results |
| --- | --- |
| Medline | 2904 |
| APA PsycINFO | 1895 |
| Ovid EMCARE | 1764 |
| HMIC | 67 |
| CINAHL | 2734 |
| Cochrane (CENTRAL) | 524 |
| **Total** | **9888** |

**Appendix 3:**
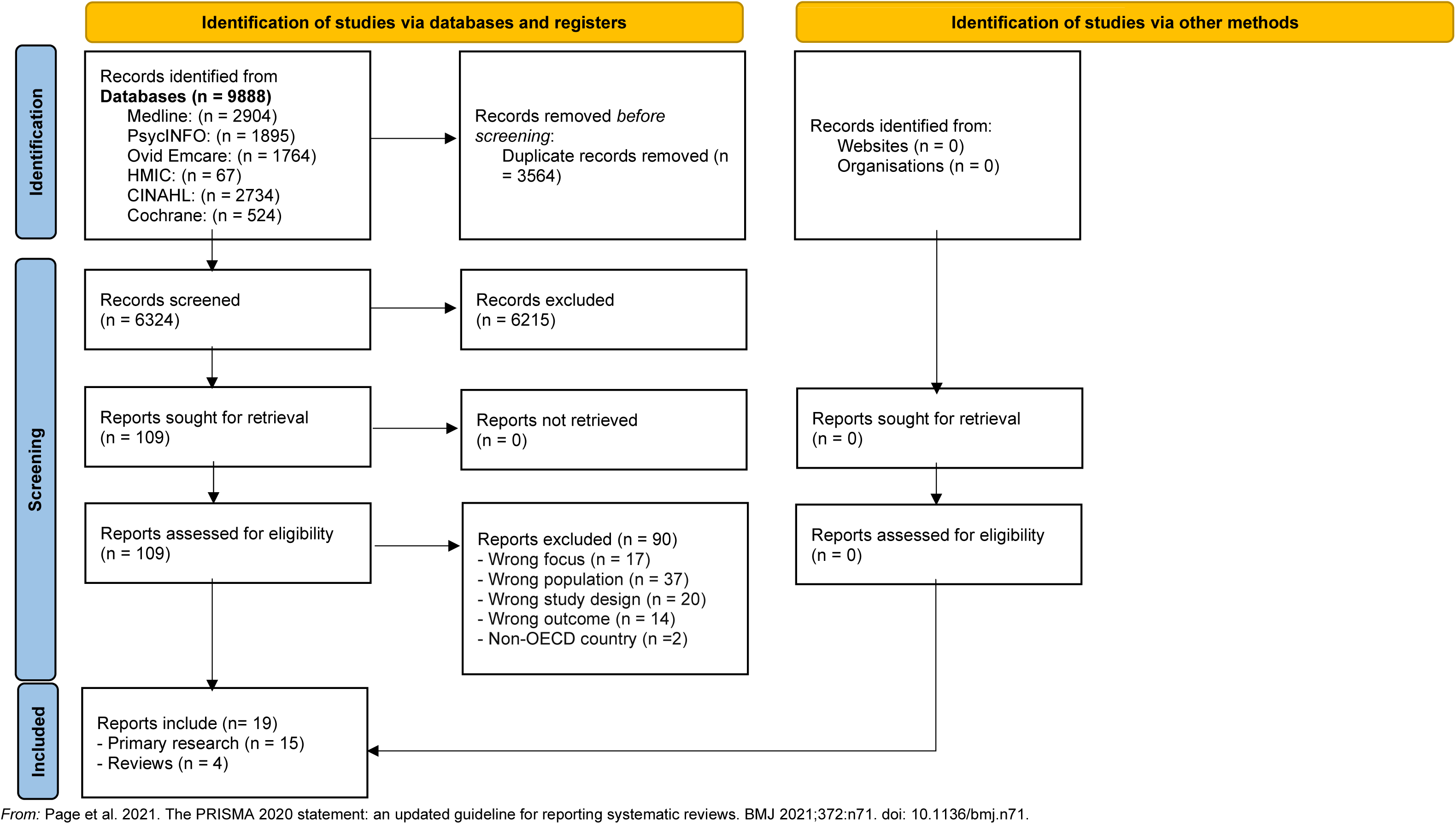
Studies excluded on full text screening.

**Appendix 3:**
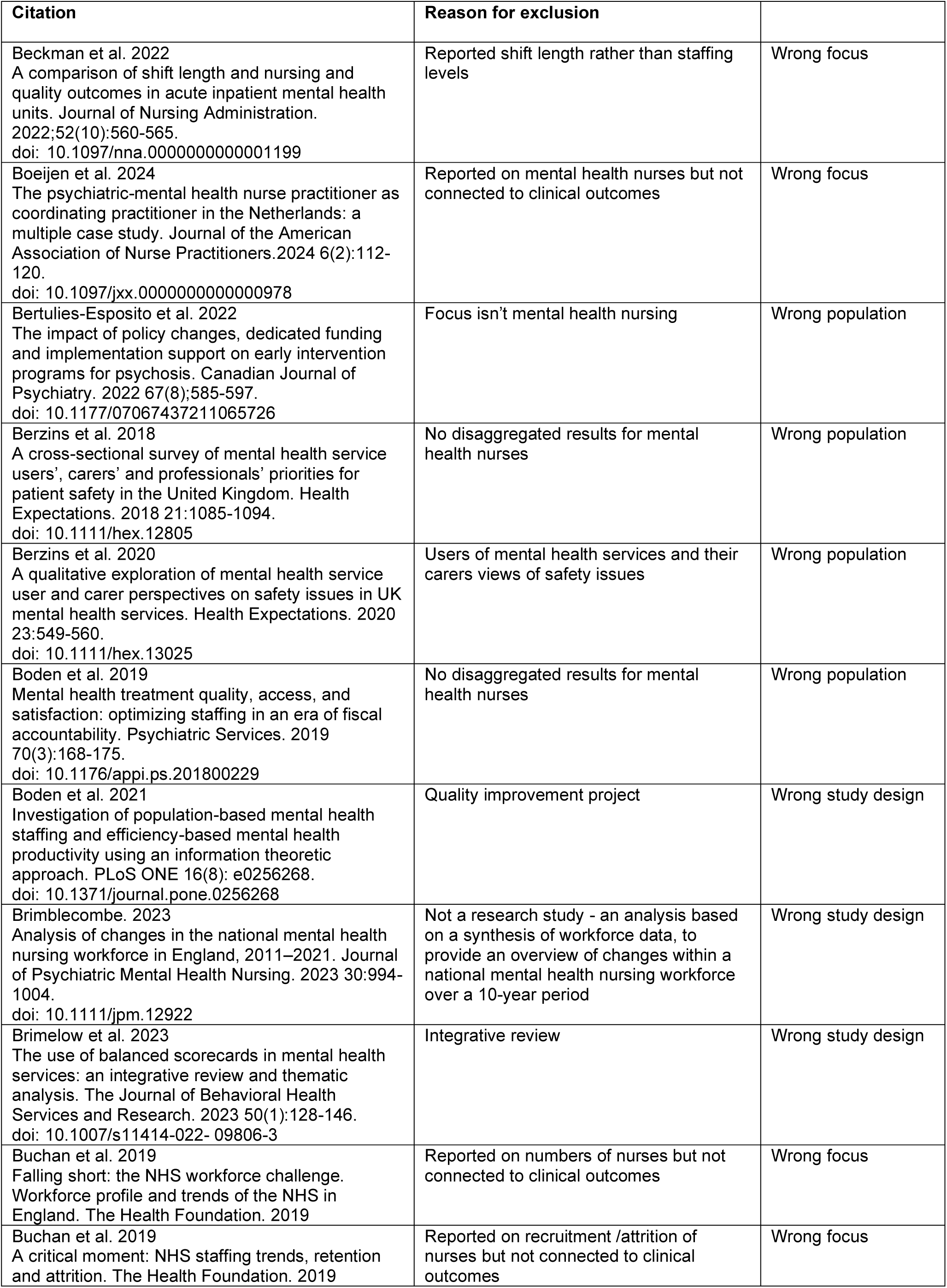
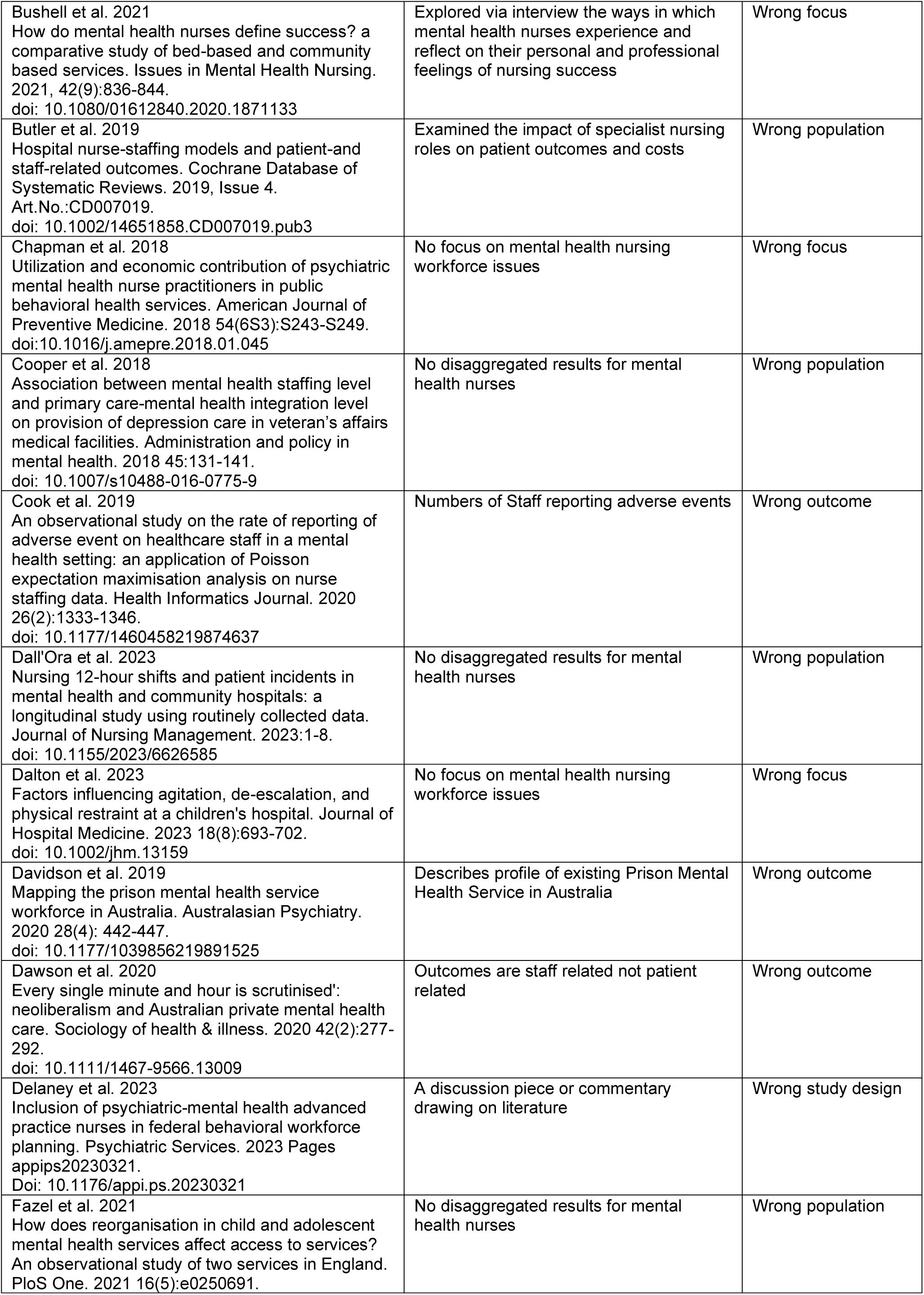
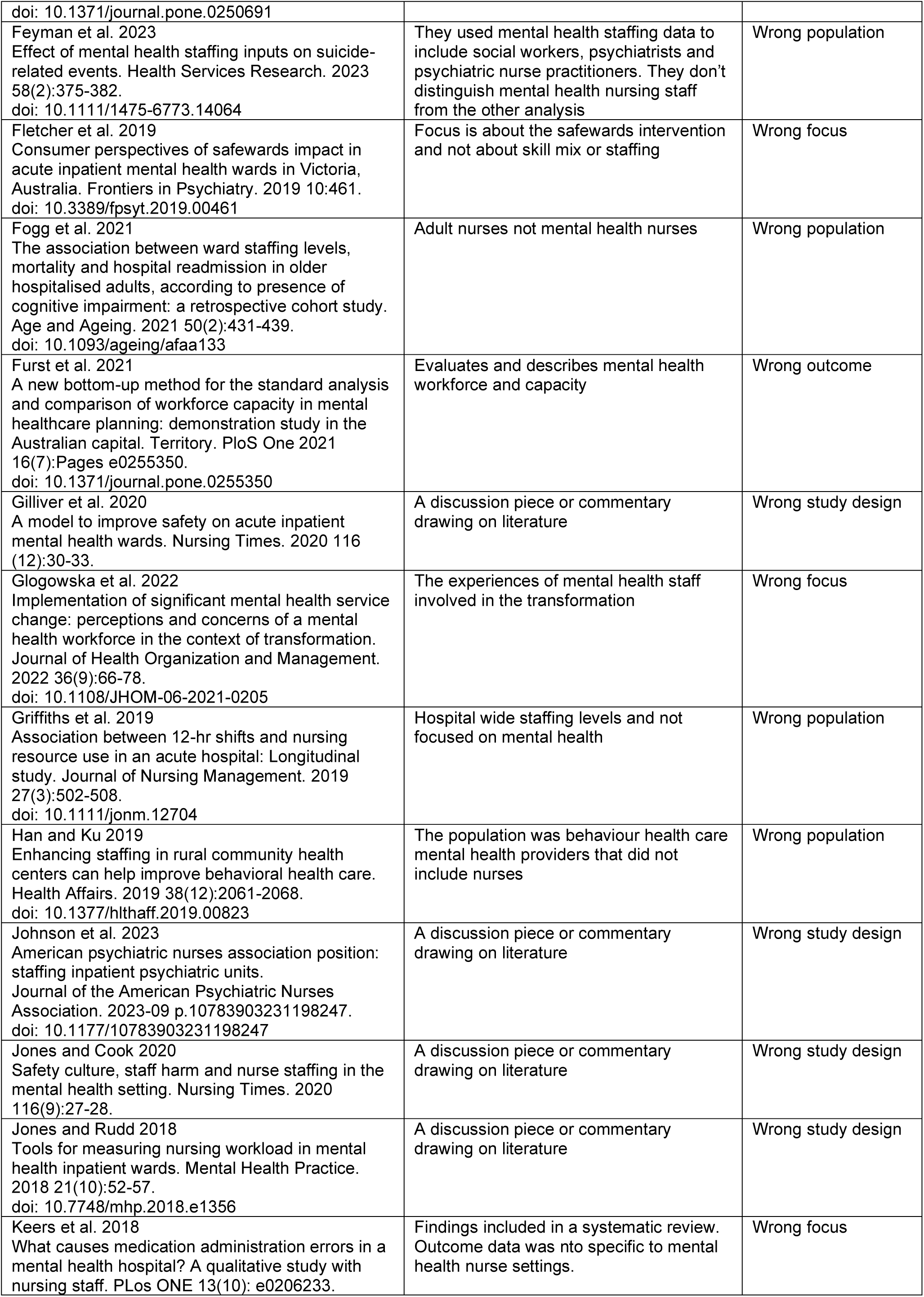
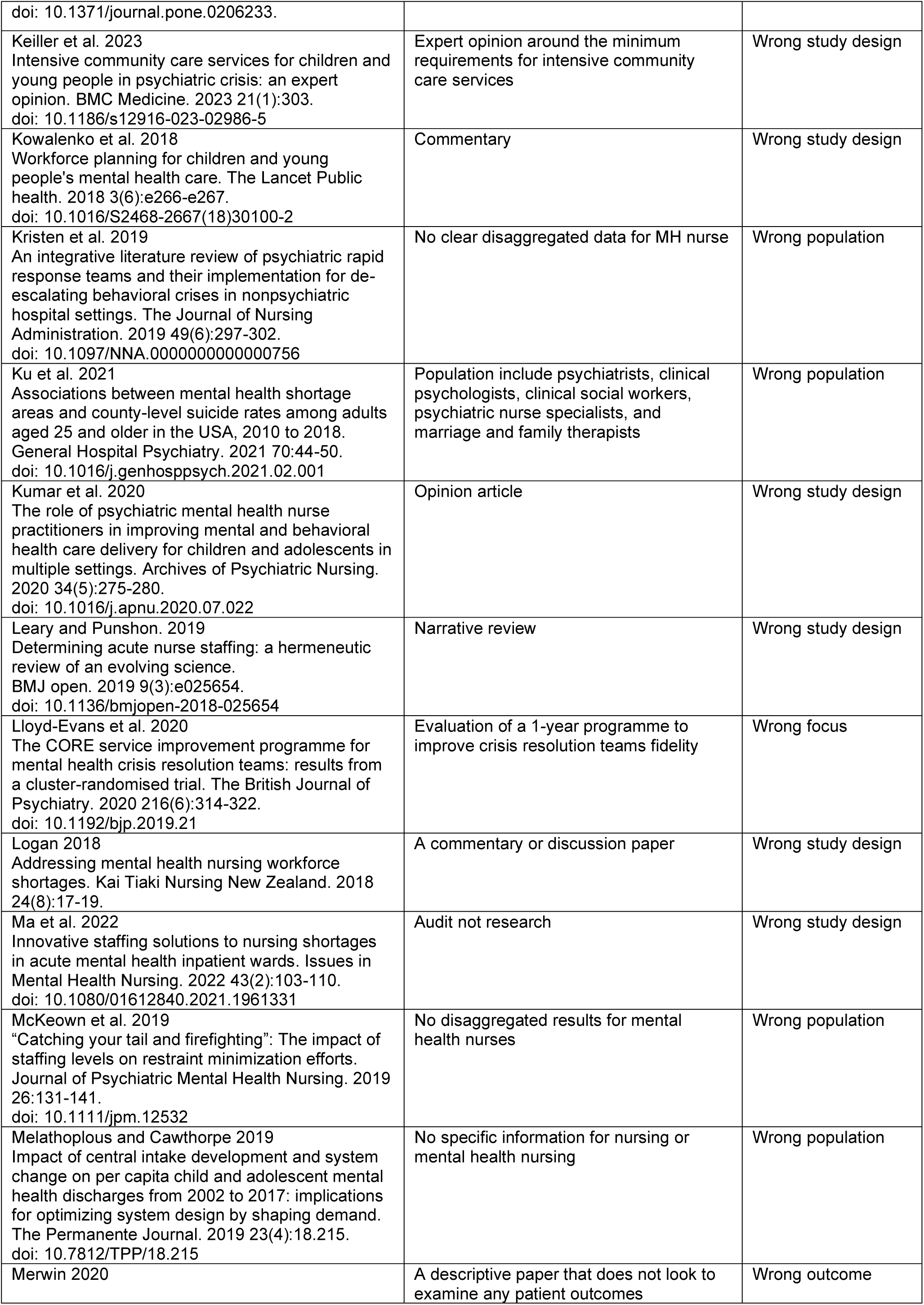
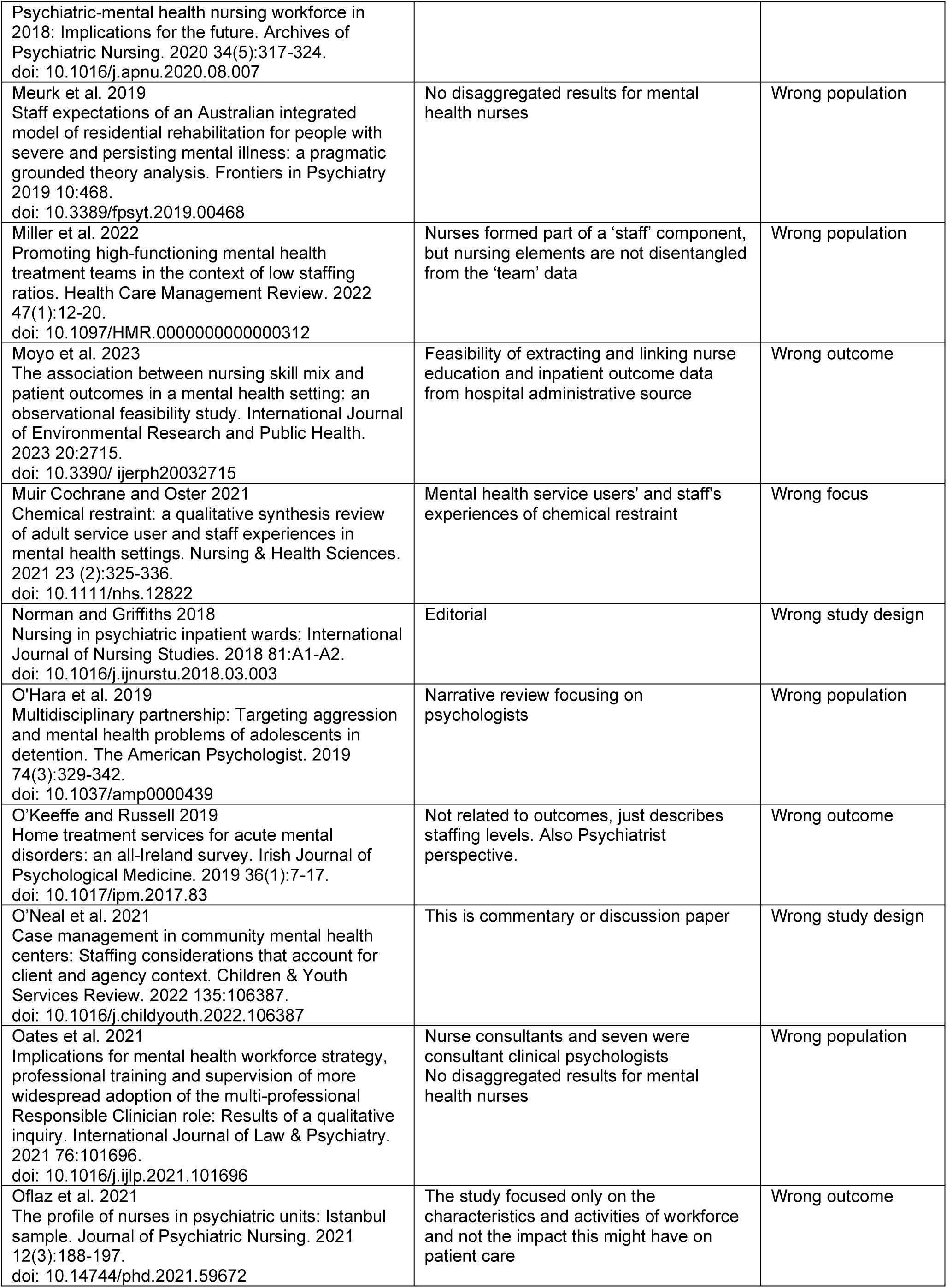
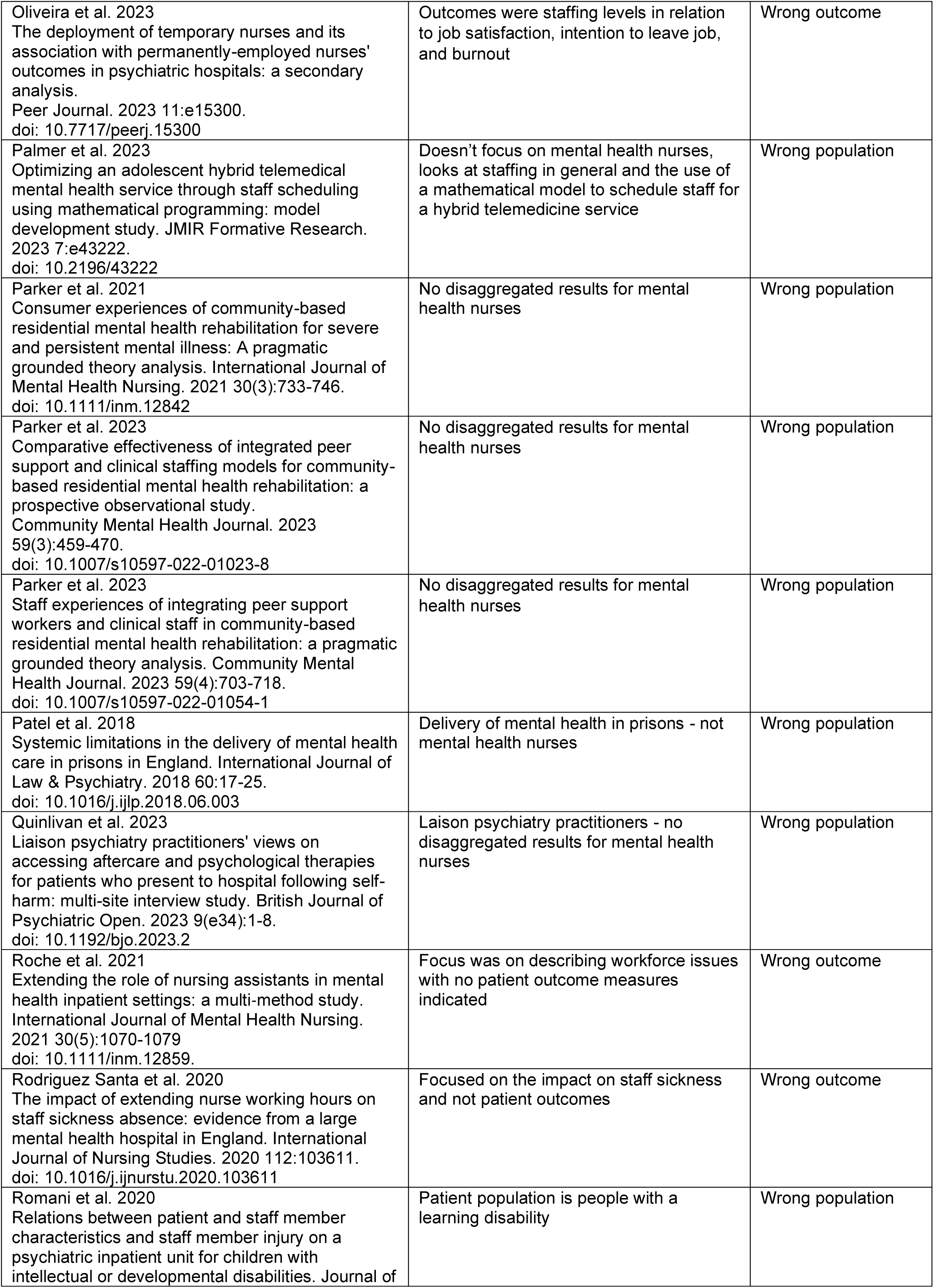
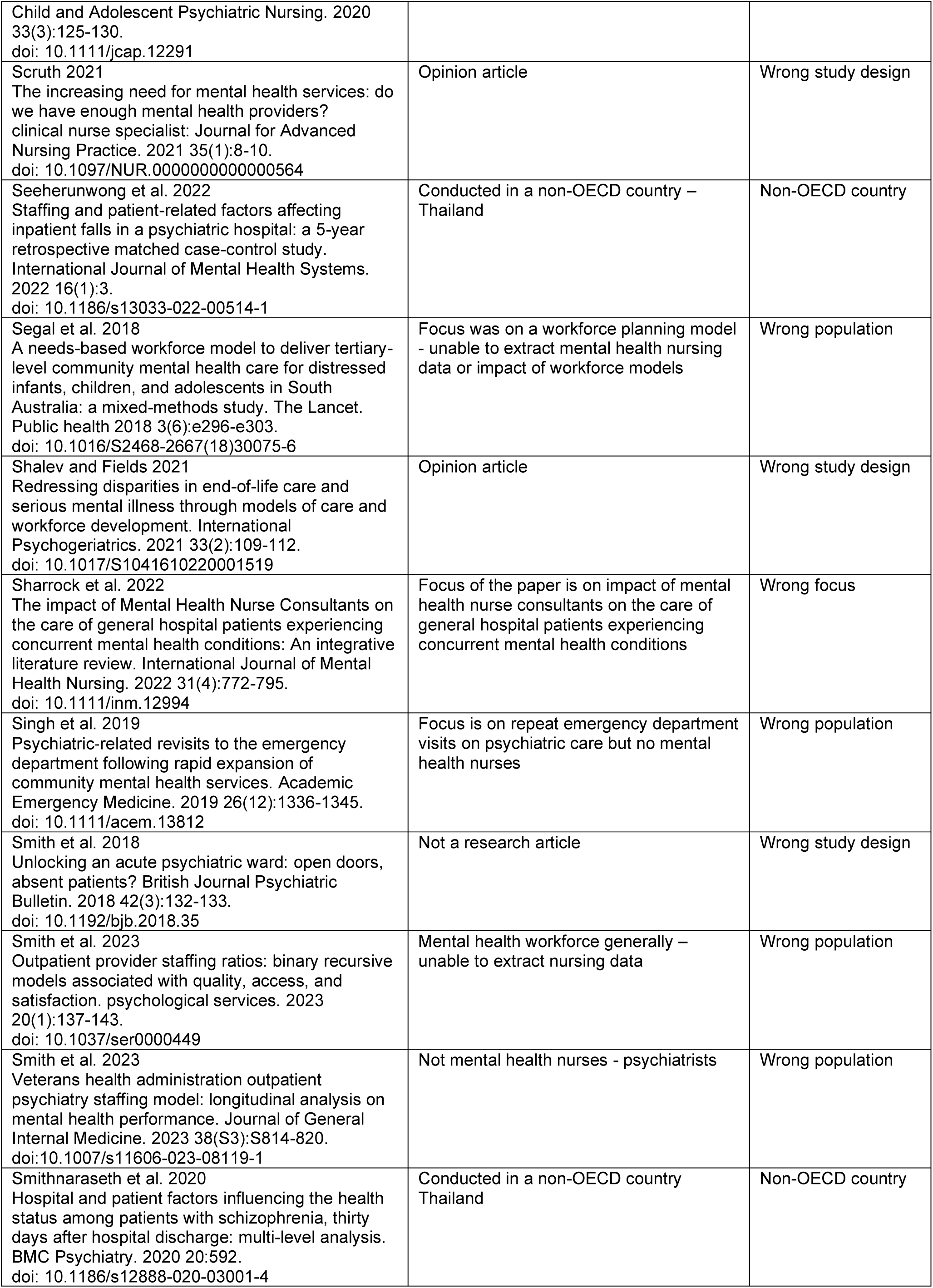
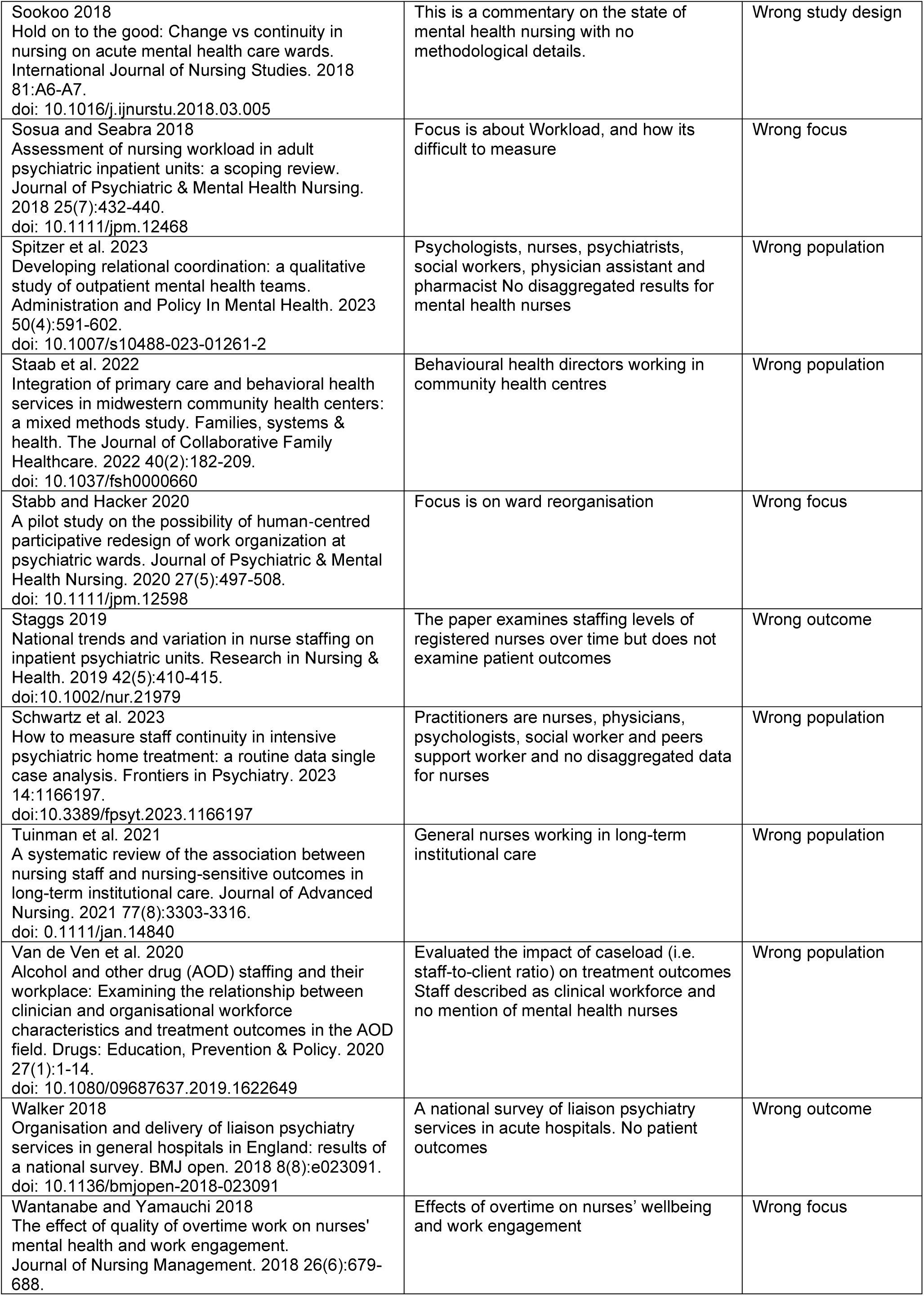
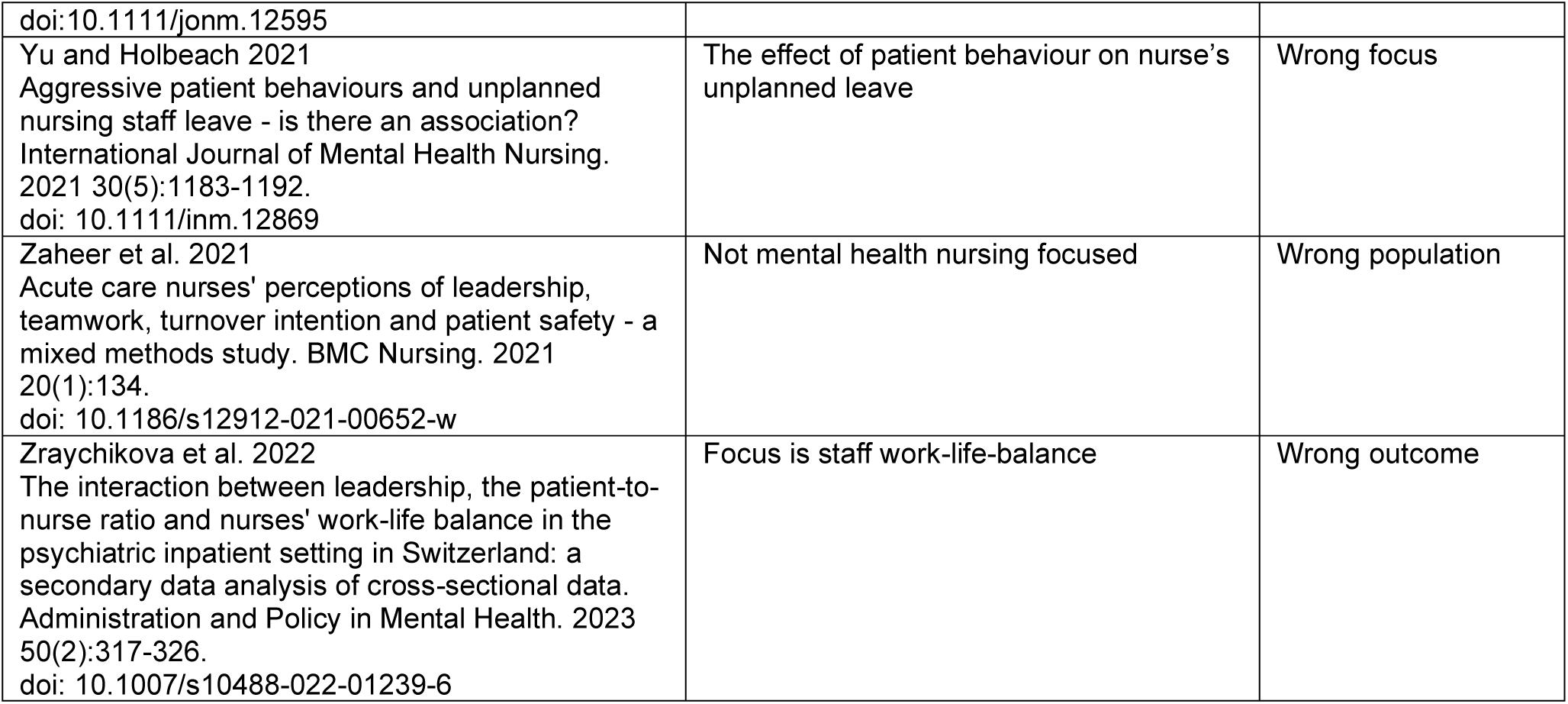
Studies excluded on full text screening.

**Appendix 4:**
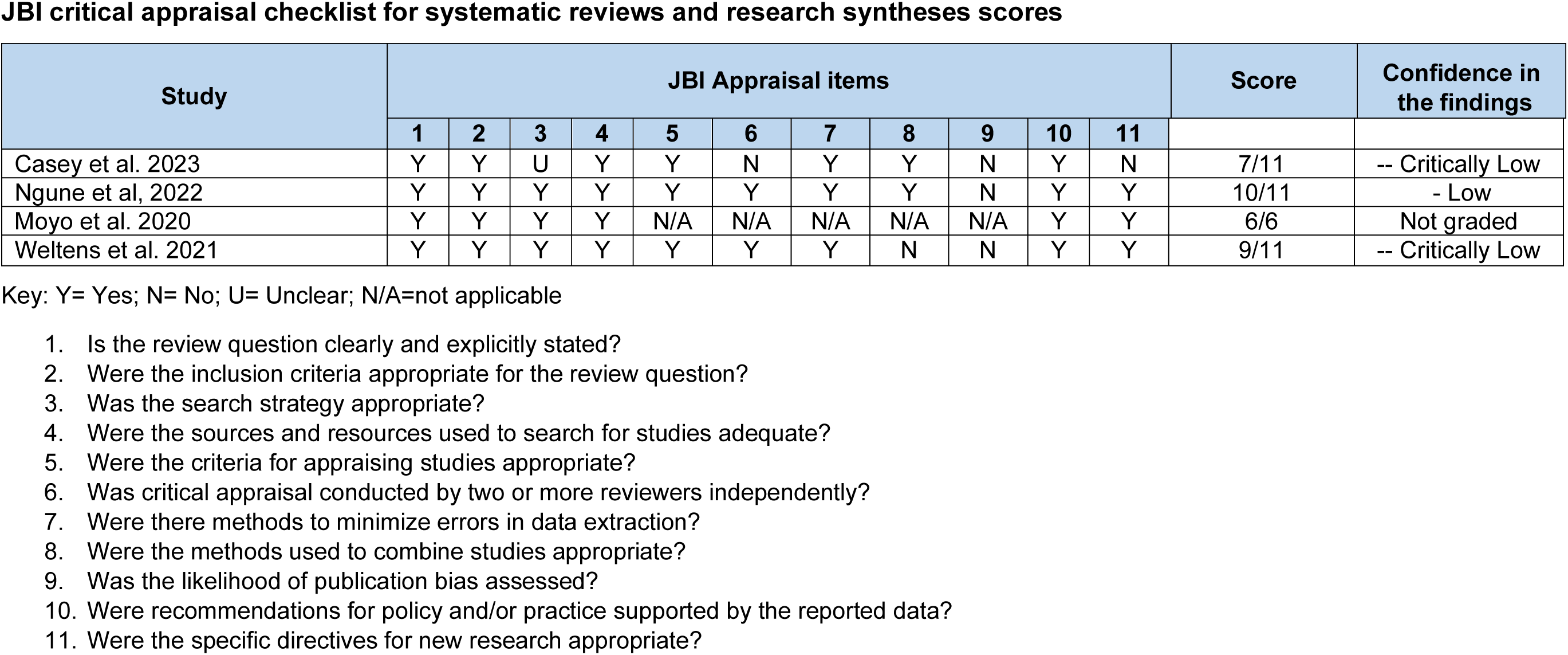
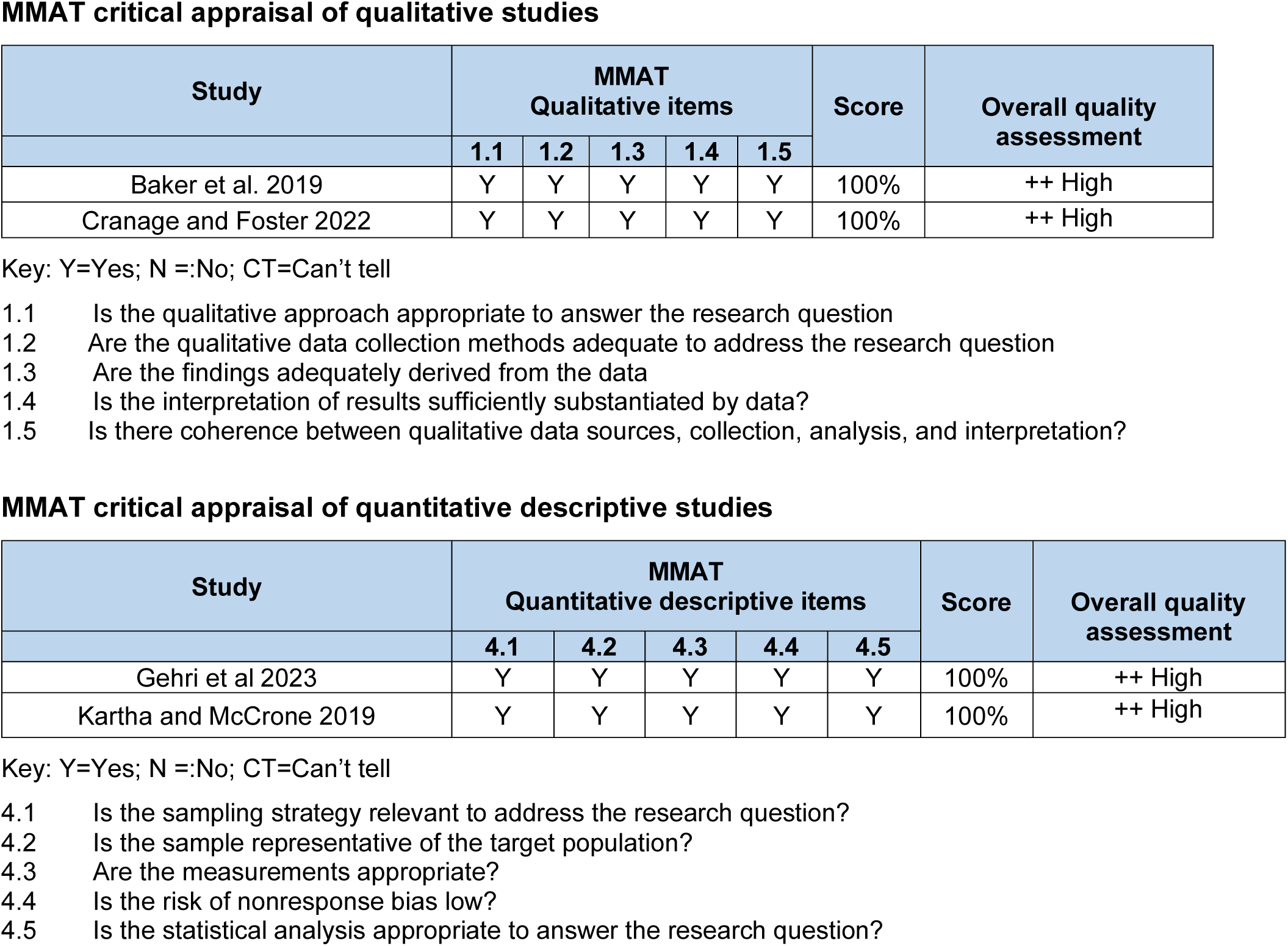
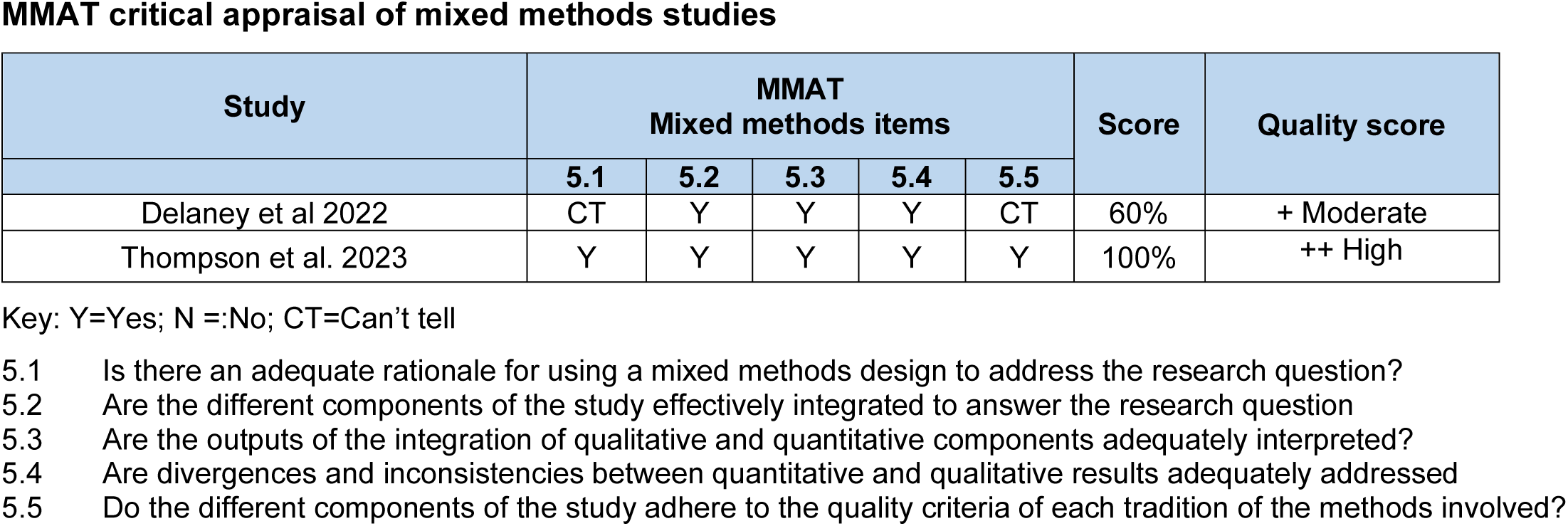
Critical appraisal scores.

## Abbreviations

Acronym: Full Description
NQB: National Quality Board
NHS: National Health Service

## Footnotes

1 Skill mix was defined as the number of registered nurses compared to other groups.

2 Skill mix was defined as the number of registered nurses compared to other groups.

3 The authors explained that the unit had a lead nurse (Senior Nurse Leader) who provided leadership and support for the whole unit, while a shift coordinator provided leadership and support for each ward within the unit.

4 Nurse types was defined as a higher proportion of registered nurses

5 Refers to a composite value that includes patient self-harm, aggression, medication, treatment or procedure, care implementation, documentation, clinical assessment and transfer.

6 Bowers, L., & Crowder, M. (2012). Nursing staff numbers and their relationship to conflict and containment rates on psychiatric wards-a cross sectional time series poisson regression study. International Journal of Nursing Studies, 49(1), 15–20.

7 Day Shifts: Low staff scenario has a staff-to-bed ratio of 1:>4; registered staff mean 2.7; non-registered staff mean 1.9. Night shifts: Low staff scenario has a staff-to-bed ratio of 1:>6; registered staff mean 1.5; non-registered staff mean 1.5.

8 Containment was defined as PRN medication, intermittent observation, constant observation, coerced intramuscular medication, show of force, manual restraint, seclusion and time out.

9 Relative technical efficiency was determined as the balance between resources (e.g. staff) and outcomes (e.g. length of stay or number of patient contacts) relative to similar services.

10 Relative technical efficiency was determined as the balance between resources (e.g. staff) and outcomes (e.g. length of stay or number of patient contacts) relative to similar services

11 Adjusted for sex and age and treatment related characteristics included psychiatric diagnosis, form of admission on the first day (voluntary versus involuntary), prescribed dose of antipsychotics at the time of admission (converted into an equivalent dose of chlorpromazine), severity of symptoms, and length of hospital stay

12 Adjusted for patient characteristics which included age, gender, type of insurance, diagnosis, previous psychiatric hospitalization within the last year, number of psychiatric sub-diagnoses, number of physical sub-diagnoses and Elixauser Comorbidity Measures score for the last year and system characteristics which included type of hospital, size, ownership, teaching, location, bed operation rate, and RN proportion (the ratio of RNs to total nursing staff).

13 Number of individuals receiving at least one antipsychotic prescription during 2015 per 1,000 inhabitants).

14 Model 1 - Adjusted for psychiatric beds (x 100,000 inhabitants); treated prevalence of mental disorders (x 100,000 inhabitants); treated incidence of mental disorders (x 100,000 inhabitants), psychiatric hospital admissions (x 100,000 inhabitants); poverty index; employment rate. Model 2 Adjusted for psychiatric beds (x 100,000 inhabitants); treated prevalence of schizophrenia (x 100,000 inhabitants); treated prevalence of bipolar disorder (x 100,000 inhabitants); treated incidence of schizophrenia (x 100,000 inhabitants); treated incidence of bipolar disorder (x 100,000 inhabitants); psychiatric hospital admissions (x 100,000 inhabitants); poverty index; employment rate.

15 Multiple non-critical weaknesses may diminish confidence in the review and it may be appropriate to move the overall appraisal down from moderate to low confidence

